# Immune profiling, microbiome, metabolomics, and gut physiology of a 1-year controlled human hookworm infection

**DOI:** 10.1101/2023.03.14.23287270

**Authors:** Francesco Vacca, Brittany Lavender, Sophia-Louise Noble, Alissa Cait, Kate Maclean, John Mamum, Bibek Yumnam, Tama Te Kawa, Thomas C Mules, Laura Ferrer-Font, Jeffry S. Tang, Olivier Gasser, Graham Le Gros, Mali Camberis, Stephen Inns

## Abstract

The observation that experimental helminth infection can be associated with immunomodulation and suppression of inflammatory diseases at distal tissue sites, has been used as rationale for trialing helminths such as *Necator americanus* for the treatment of inflammatory disorders in humans. However, the lack of sufficient knowledge of the immunological interplay between human host and parasite in a controlled infection setting limits ongoing clinical intervention studies. In this one-year longitudinal study, healthy volunteers were recruited and infected with *N. americanus*. Changes in immune responses, microbiome, plasma metabolome and gut physiology were examined over the course of the one-year period. All participants were successfully infected as confirmed by detectable eggs in the feces and adult worms visualized in the intestine. In general, individual variation in immune cells, serum cytokines, fecal microbiome and plasma metabolites were greater than changes induced by the infection. Nevertheless, eosinophils, serum IL-5, and fecal eosinophil degranulation markers transiently increased in the acute phase of infection. In addition, while we observed stability in microbial community composition through the course of infection, we found a difference in the microbial community composition of participants with moderate gastrointestinal symptoms. No significant changes were observed in gut physiology measured using Smartpill^TM^, except for a decrease in small bowel pH. Untargeted plasma metabolomics analysis of participant plasma over the course of infection revealed enrichment in tryptophan metabolism following infection which was associated with increased CTLA-4 expression on regulatory T cells (T_REGS_), CRTH2^+^ T helper 2 cells (T_H_2) and CCR6^+^ T helper 9 cells (T_H_9). In conclusion, hookworm infection is well tolerated and represents an innovative platform for investigating immunomodulatory properties of hookworm infection in a therapeutic clinical setting.

**One Sentence Summary:** Controlled human hookworm infection changes immune-linked metabolic pathways

## INTRODUCTION

*Necator americanus* (*Na*) is a species of hookworm that infects many millions of people in subtropical areas of the world(*1–3*). The parasite/host relationship that has evolved between *Na* and humans has been formed over several millions of years (*4*) and features a immunological accommodation by the human host of the adult worm for up to ten years. The long term nature of hookworm infection of humans implies that compromising host survival is not in the best interests of the hookworm, and it has been hypothesized that chronic *Na* infection can confer beneficial immunomodulatory effects on the human host (*5, 6*). This and the observation that inflammatory diseases have become increasingly prevalent in regions where hookworm has been eradicated, has led researchers to investigate potential positive effects of hookworm infection(*6, 7*). Pioneering studies carried out in Africa and South America have observed reductions in atopic sensitization in those individuals carrying hookworm (*8–10*). Furthermore, populations from hookworm endemic areas present with a signature regulatory T cell (T_REG_) phenotype that is not found in non-infected populations. Anti-helminth treatment decreased this regulatory T cell signature, providing further evidence for helminths ability to regulate the immune system of their human host(*11*). Clinical studies aimed at defining the immunoregulatory effects of hookworm infections in healthy volunteers demonstrate transient production of type 2 cytokines and acute increases in eosinophilia, that return to background levels as the adult hookworm establishes itself in the small intestine, indicating an active immune suppression of the human host by the adult parasites(*12–15*). Further clinical studies have determined the safety profiles of *Na* infection, immune and immunoregulatory responses, for vaccine development, and patients suffering from IBD, celiac disease and multiple sclerosis (*12, 16–24*). We sought to extend these controlled hookworm infection studies, by investigating the effect of the different phases of a controlled hookworm infection on the immune regulatory networks, microbiome, metabolome, and gut physiology of healthy individuals. Participants were followed for one year and 30 *Na* L3 infective larvae were used to establish stable infection. The potency of infection was determined by collecting stool samples at multiple time points for egg counts. All participants were successfully infected with *Na* and a transient increase in eosinophils and markers associated with eosinophilic inflammation was observed. No other serum cytokines or immune cells were perturbated by the hookworm infection. While microbial communities remained stable during the infection, participants who experienced moderate gastrointestinal symptomology had a significantly different microbiome from those who experienced mild or absent symptoms. We also observed a decrease in small intestine pH. Furthermore, hookworm infection-induced changes were observed for tryptophan metabolism and expression of CTLA-4 in T_H_2, T_REGS_ and T_H_9 cells. With this study we have been able to gain a comprehensive picture of the range of human immunological, metabolic, and physiological responses induced by *Na* infection in healthy volunteers. This data set provides a platform for considering the use of controlled hookworm infection for the treatment of inflammatory diseases and vaccine development in humans.

## RESULTS

### *Necator americanus* infection induces tissue-specific symptoms

In this study 13 individuals with no history of allergies, auto-immune disease or inflammatory gut conditions completed the study (Fig1A and Table 1). We inoculated participants with 30 infective L_3_ (iL_3_) and followed host immune responses, microbiome, and gut physiology (Fig. 1B) during the acute and chronic phase of infection (Fig. 2A). Participants monitored symptoms they experienced for the first 12 weeks of infection (e.g., dermatological reaction, respiratory and gastrointestinal symptoms). Skin reactions (e.g., rash and itchiness) were observed mainly during the first weeks (Fig. 2B). Similarly, minor respiratory symptoms (e.g., cough, sneezing or sore throat) were observed by one participant during the first weeks (Fig. 2B), consistent with the lung phase of the parasite infection cycle. The majority of gastrointestinal (GI) symptoms were reported at 5- and 6-weeks post-infection, indicating the parasite reaching the gut lumen (Fig. 2B). No severe reactions were reported, indicating good tolerability of *Na* infection. With a view to enabling long-term storage and batched supply of hookworm larvae, cryopreservation was performed using methods previously described(*25*) and 4 of the 12 participants of this study received cryopreserved larvae which had been reanimated. Eggs were detectable in fecal samples from week 8 of infection onwards in all participants with similar egg production kinetics observed for those participants that received either cryopreserved or fresh larvae (Fig. 2C). Transient GI symptoms preceded egg detection in the feces (Fig. S1A) and two participants that chose to retain the hookworms displayed continued egg production for over 2 years post-infection (Fig. S1B). To further confirm infection, PillCam^TM^ capsule endoscopy was performed at week 20 to visualize and enumerate adult worms at the site of infection. Adult parasites were detectable in all participants, with no differences from participants that received cryopreserved larvae (Fig. 2D), and in Fig. 1E representative PillCam^TM^ images are shown. Eosinophilia was observed as early as 4 weeks post-infection, with peak numbers in the blood occurring by 8 weeks post-infection (Fig. 2F). No correlation was observed between eggs and visualized adult worms (Fig. S1C). No differences were observed in eosinophilia between participants infected with fresh or cryopreserved larvae (Fig. 2F). Given no differences were observed in eggs counts, worm visualization and eosinophil kinetics using cryopreserved larvae, all further analyses were carried out as a combined set.

**Figure 1:**
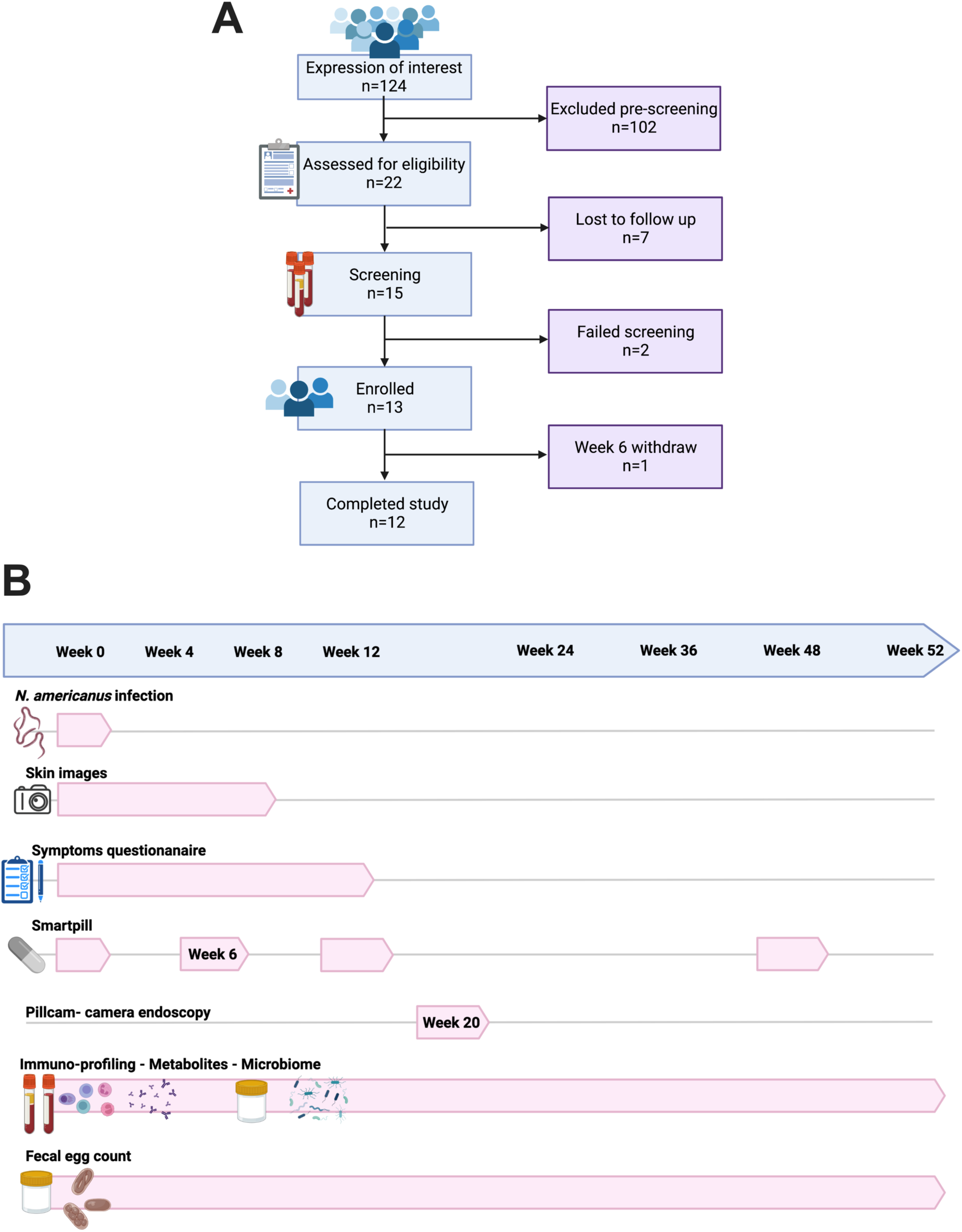
Trial profile and study design. (A) Schematic representation of the recruitment and eligibility process. Eligibility criteria described in Data File S1. (B) Study design with sample collection. Created with BioRender.com

**Figure 2:**
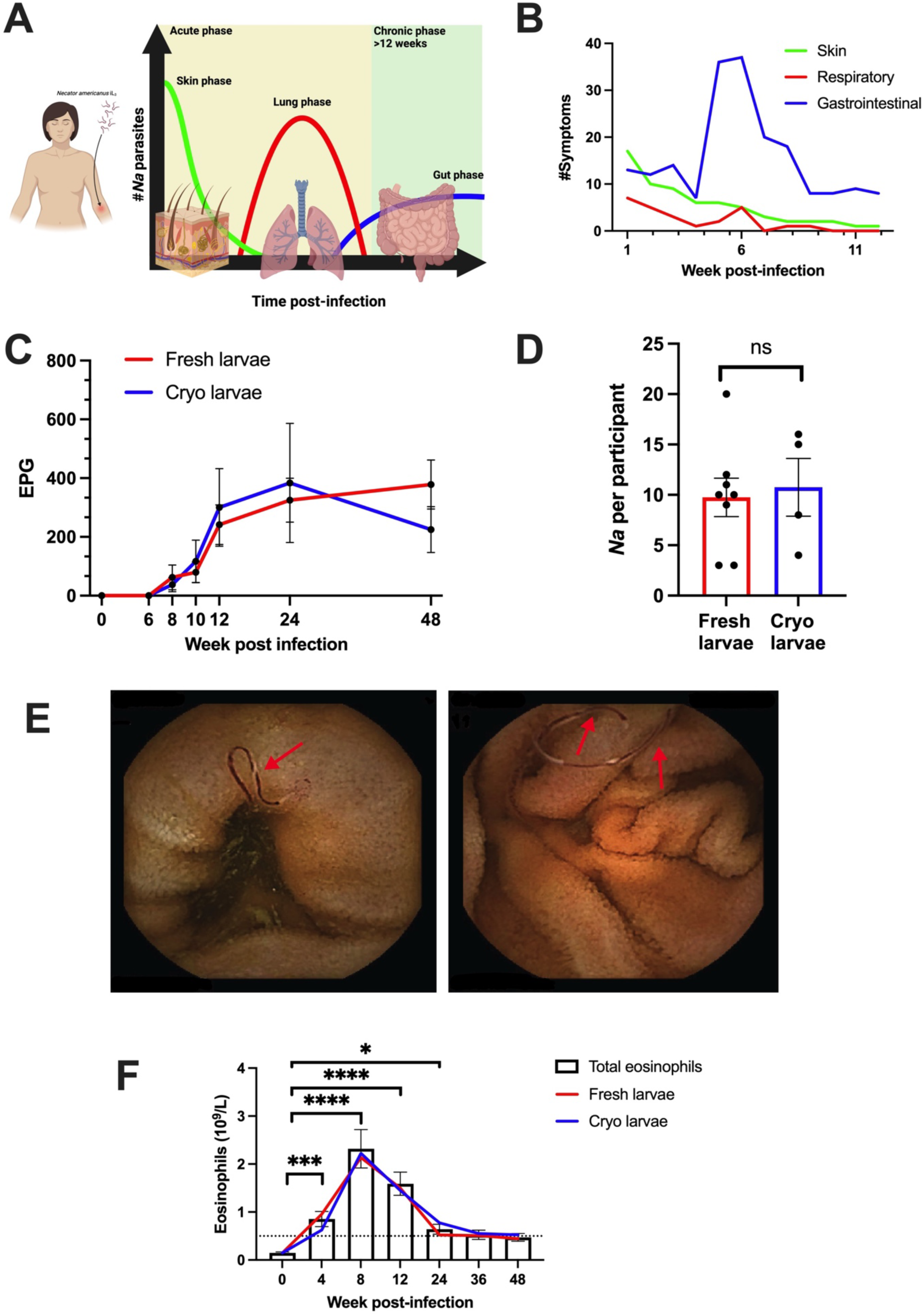
*Necator americanus* infection induces site-specific symptoms and eosinophilia. (A) Schematic representation of phases during hookworm infection investigated in this study. Created with BioRender.com (B) Self-reported symptoms divided by organs affected by parasite traverse in the host. (C) Plot showing mean ± SEM eggs count measured from fresh stool samples divided by participants that received fresh (red line) or cryopreserved (blue line) larvae. (D) Graph showing adult hookworm counted in the gastrointestinal tract with Pillcam^TM^ endoscopy divided by participants that received fresh (red) and cryopreserved larvae (blue). Showed is average from two independent counting. (E) Representative images of Pillcam^TM^ endoscopy showing hookworm (red arrow) in the intestinal tract. (F) Bar graph showing mean ± SEM of peripheral blood eosinophil count. Lines showing eosinophils count in participants infected with fresh (red line) or cryopreserved (blue line) larvae. Results were analysed with One-way ANOVA using Friedman test with Dunn’s multiple comparison test. p=*<0.05, **<0.005,****<0.0001

### Hookworm infection increases serum IL-5 and fecal eosinophil-specific degranulation markers

In our study we observed blood eosinophilia in participants infected with *Na,* with peak eosinophilia at week 8 post-infection (Fig. 2F). We analyzed IL-5 levels in serum samples collected over the course of infection and significantly increased serum IL-5 was observed up to 24 weeks post-infection (Fig. 3A), with peak levels observed at 4 weeks post-infection. There was a significant correlation between IL-5 levels and eosinophil count when comparing serum IL-5 at week 4 (preceding the eosinophil peak) and eosinophilia at week 8 (Fig. 3B), however total IL-5 levels did not correlate with total blood eosinophilia (Fig. S2A). No differences were observed in eotaxin (Fig. S2G), a chemokine involved in eosinophil recruitment. We also followed levels eosinophilic cationic protein (ECP) and eosinophil-derived neurotoxin (EDN)(*26*) in fecal samples as a readout of eosinophil activation and degranulation in the intestinal tract. An increase in both ECP and EDN was observed from week 4, with levels peaking at week 8 post-infection (Fig. 3C, 3E). Levels of ECP and EDN positively correlated with blood eosinophil count (Fig. 3D-F), but not with serum IL-5 (Fig. S2B-C). Another inflammatory marker used routinely in a clinical setting is fecal calprotectin (fCal) which is normally exclusively associated with neutrophilic inflammation(*27*). No differences were observed in blood neutrophils during infection (Fig. 3G) however we did observe a significant transient increase in fCal during the acute phase of infection with levels remaining within the normal range (Fig. 3G). Interestingly, we did not detect any increase in human neutrophil lipocalin (HNL) (Fig. 3G), a specific neutrophil degranulation marker, indicating that eosinophils might be the major inflammatory cell in the intestine during hookworm infection. Studying cytokine levels in serum, we noticed that individual variation in immune markers is greater than changes induced by the hookworm infection as shown by the PCA (Fig. S2D) and subsequently no significant differences were observed in cytokines involved in type 2 immune responses (Fig. S2E), type 1 immune responses (Fig. S2F), chemokines (Fig. S2G), and serum antibodies (Fig. S2H) when compared to baseline levels. The apparent increase in IL-17A (Fig. S2F) was specific to a participant that developed low eosinophil response upon infection. We also observed an increase in *Na*-specific IgG that reached significance at week 24 post infection (Fig. S2K). No differences were observed when measuring the known immunomodulatory cytokines TGF-β1(Fig. S2I) and IL-10 (Fig. 2SJ). Taken together, these data indicate that infection with 30 iL3 induces a transient increase in IL-5 cytokine levels that is followed by a significant increase in eosinophilia.

**Figure 3:**
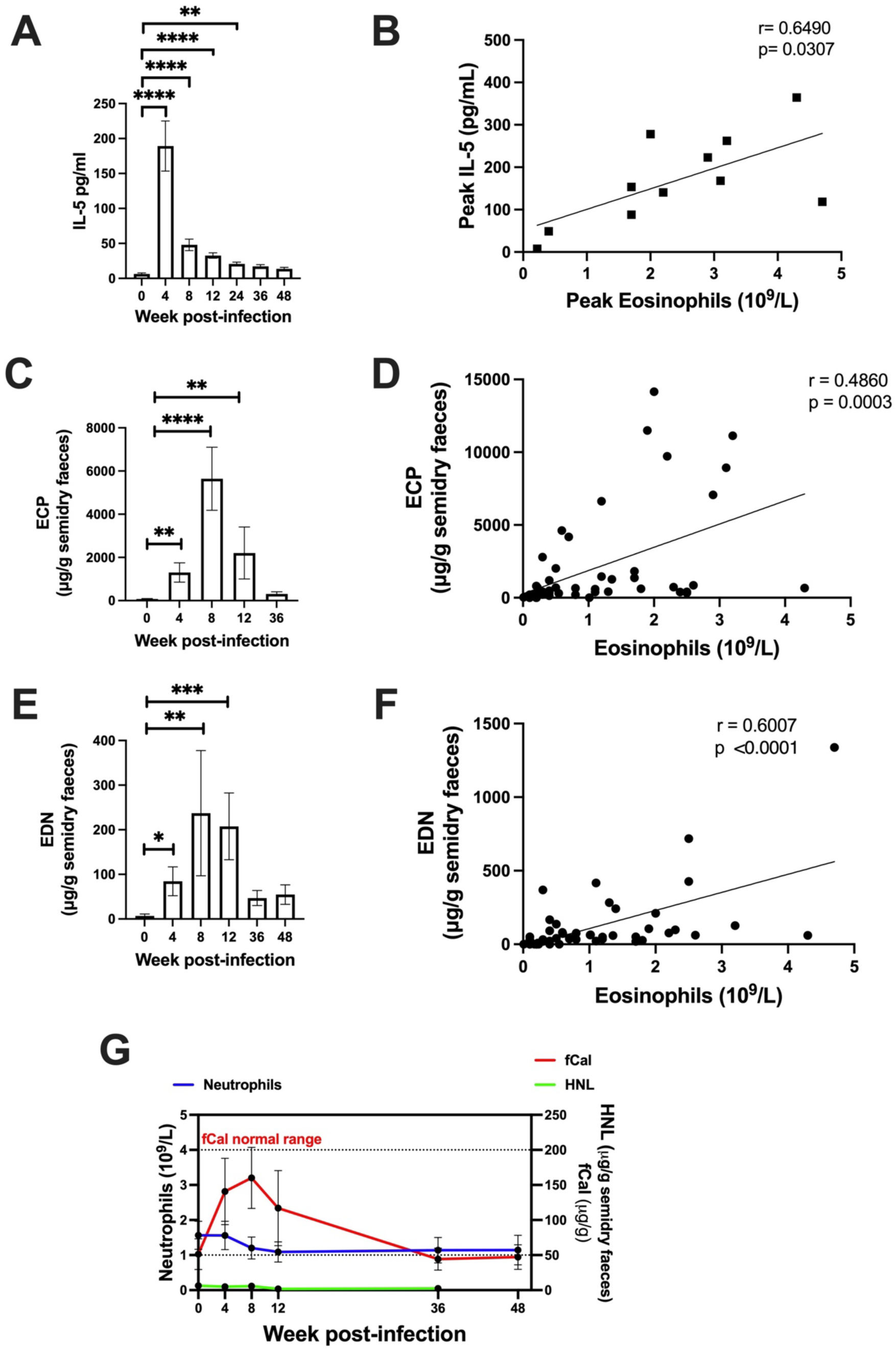
Hookworm infection increases serum IL-5 and faecal eosinophil-specific degranulation markers. (A) Bar graph showing serum IL-5 mean ± SEM. (B) Scatter plot showing correlation between peak serum IL-5 (week 4) and peak eosinophils count (week 8). (C) Bar graph showing ECP mean ± SEM measured in faeces. (D) Correlation between ECP levels and eosinophils count over the course of infection. (E) Bar graph showing EDN mean ± SEM measured in faecal samples. (F) Correlation between EDN levels and eosinophils count over the course of infection. (G) Faecal calprotectin (fCal – red line) plotted against neutrophils count (blue line) and HNL (green line). Showing mean ± SEM. (A) Analysed with One-way ANOVA using Friedman test with Dunn’s multiple comparison test. (B, D and F) Analysed with Pearson correlation. (C and E) Analysed using One-way ANOVA using Kruskal-Wallis with Dunn’s multiple comparison test. p=*<0.05,**<0.005,***<0.001,****<0.0001.

### Peripheral blood immune cells are unchanged following *N. americanus* infection

We analyzed peripheral blood mononuclear cells (PBMCs) by high dimensional flow cytometry and following an unbiased High Dimensional Analysis approach. Normalization was performed to remove batch to batch variation, we then ran FlowSOM algorithm for clustering the cells and used Uniform Manifold Approximation and Projection (UMAP) to reduce the dimension of the data and to visualize the immune cells in maps. Two panels were used to analyze PBMCs (Fig. S3A-B). According to their receptor expression, we were able to identify CD4^+^ and CD8^+^ T cells, γδT cells, T_REG_, MAIT cells and NKT cells for panel 1 (Fig. 4A), and B cells, dendritic cells, NK cells and monocytes for panel 2 (Fig. 4D). We wanted to investigate if hookworm infection induced perturbation in immune cells apart from eosinophils. For both panel 1 and panel 2, density plots were generated to visualize any possible change in the main immune cell populations (Fig. 4B and E). We observed a high degree of variability amongst participants, however no significant differences were observed in immune cell frequency (Fig. 4C and F). In general, immune cell variability between individuals was greater than changes induced by the hookworm (Fig. S4A). The two panels allowed us to study several sub-populations in the CD4^+^ T cell, CD8^+^ T cell, B cell and DC populations as shown in the gating strategies (Fig. S3A-B), accordingly to markers expression (Fig. S4B and E), and we then overlapped subpopulation using UMAPs in Fig. 4 (Fig. S4C and F). From all sub-populations analyzed no significant differences were observed after hookworm infection as shown by the heat maps (Fig. S4D and H). Taken together these results suggest that infection is well tolerated, and no abnormal immune responses were observed.

**Figure 4:**
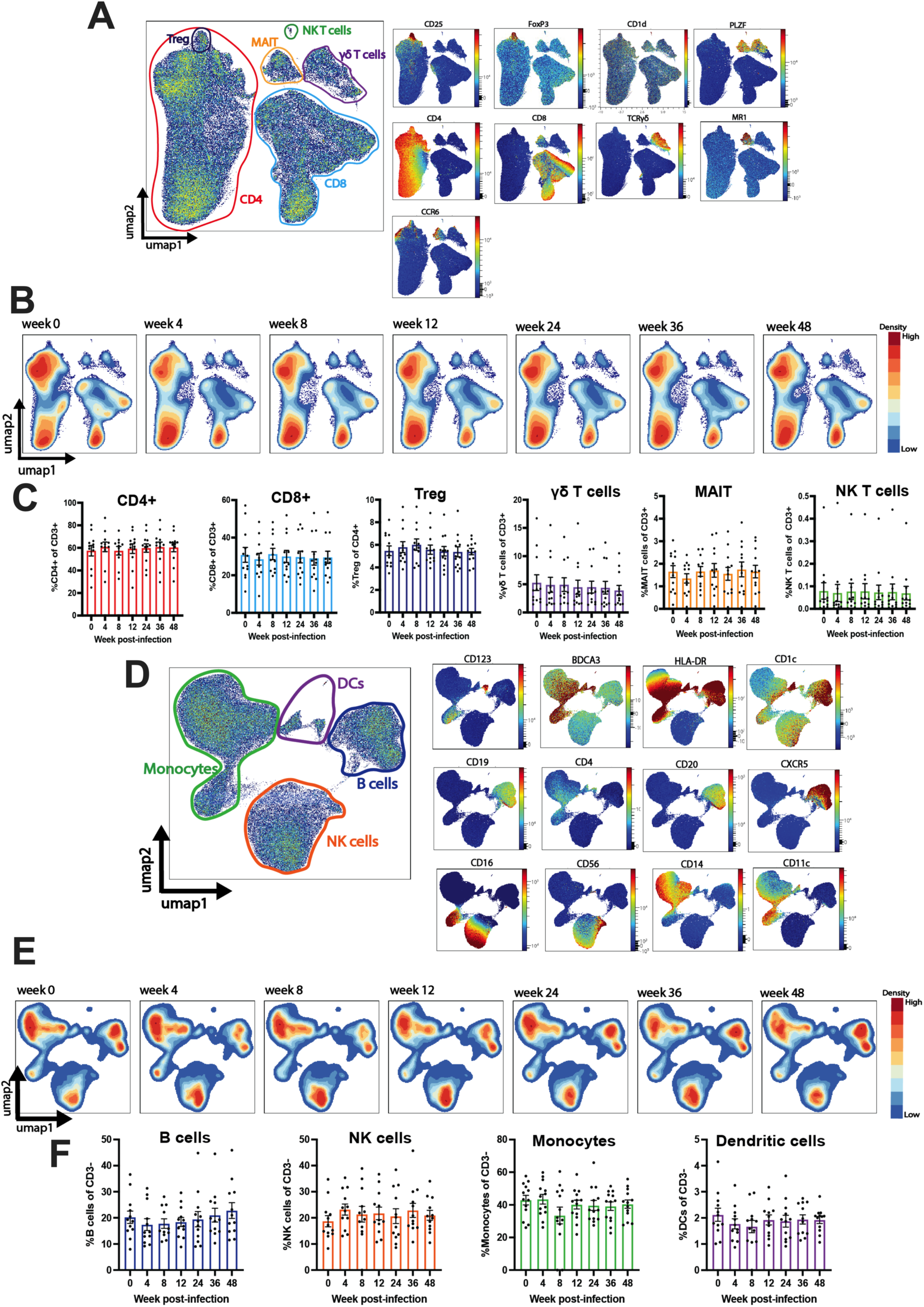
Peripheral blood immune cells are unchanged following *N. americanus* infection. (A)Panel 1 UMAPs identifying main populations and representative marker expression. (B) Panel 1 UMAPs showing population density over the course of infection. (C) Bar graphs showing individual values and mean ± SEM for immune cell population over time. (D) Panel 2 UMAPs identifying main populations and representative marker expression. (E) UMAPs showing population density over the course of infection. (F) Bar graphs showing individual values and mean ± SEM for immune cell population over time for panel 2.

### Microbiome stability and gut physiology during hookworm infection

We examined fecal samples for microbiome analysis at the indicated time points during infection with *Na*. We visualized beta-diversity by principal component analysis (PCA; Fig 5A) and found inter-individual microbiome composition was the greatest source of variation in the data and no significant changes to beta-diversity induced by hookworm infection (PERMANOVA, P>0.1) Microbial community stability (Bray-Curtis dissimilarity relative to D0) (Fig. 5B) and alpha diversity (Shannon diversity index) (Fig. 5C) were not significantly different at any sampled timepoint post infection. Interestingly, participants that experienced moderate GI symptomology (Fig. 5D, showed in red) had a significantly different microbiome from those who experienced mild symptoms or were symptom free (shown in dark blue; PERMANOVA P < 0.001) (Fig. 5D), had higher alpha-diversity (Fig. 5E), and a significantly higher relative abundance of the phylum Firmicutes (Fig. 5F, Fig. S5A, Table S2). At the OTU level we identified 130 taxa that were differentially abundant in participants who experienced moderate GI symptoms (Fig. S5B, Table S3). We observed a linear correlation between fCal levels and alpha diversity (Fig. S5C) and several OTUs with highly significant linear correlations with fCal levels, particularly from the phylum Firmicutes (Fig. S5D). Participants that experienced moderate symptoms did not display higher eosinophil levels at week 4 (Fig. S5E), but they had higher fCal (Fig. S5F), and a trend towards increased ECP and EDN in the feces (Fig. S5G and H respectively). Furthermore, we examined gut motility and physiology using Smartpill^TM^. We observed a trend towards reduced intestinal transit time at 12 weeks post infection (Fig. S5I) and reduced stomach and small bowel contractility at week 12 (Fig. S5J-K). We found no difference in colon contractility (Fig. S5L). However, we did observe a pH decrease in the small intestine 6-weeks post-infection (Fig. 5G). No significant differences were observed in stomach and colon pH (Fig. S5M-N), and pressure in the stomach and small bowel (Fig. S5O-P).

**Figure 5:**
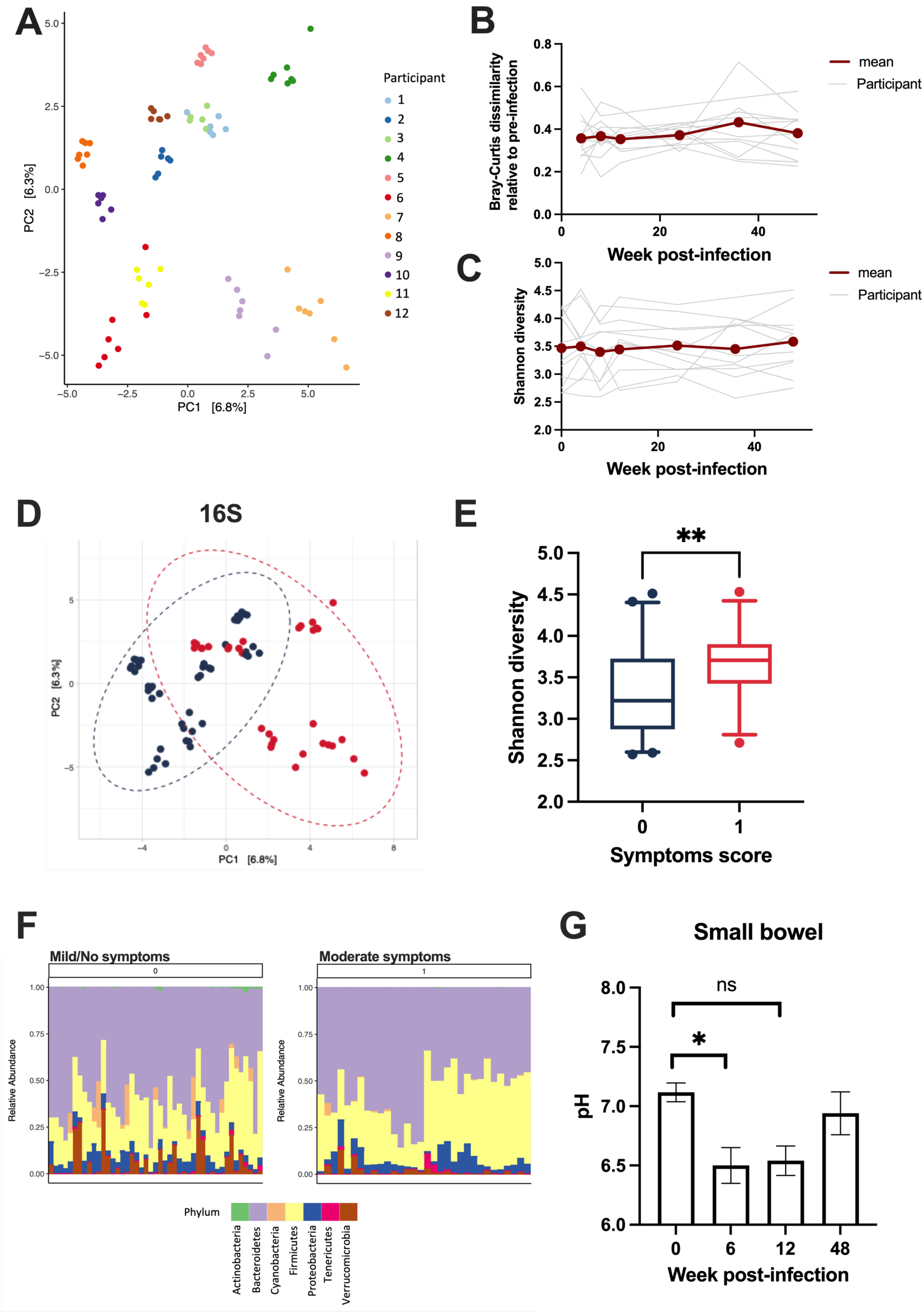
Microbiome stability and small bowel pH during hookworm infection. (A) PCA plots showing individual participants (B) Graph showing Bray-Curtis dissimilarity test for each single participant (grey lines) and mean (dark red line). (C) Shannon diversity showing each participant (grey lines) and mean (dark red line). (D) PCA plot showing 16s analysis grouping participants according to symptoms score (no/mild symptoms (dark blue); moderate symptoms(red)). (E) Box plot showing Shannon diversity accordingly to symptoms (score 0= no/mild symptoms (dark blue); 1= moderate symptoms(red)). (F) Phylum relative abundance for participants with mild/no symptoms (0) and moderate symptoms (1). (G) Small bowel pH measured by Smartpill. Showing mean ± SEM (N=6). (E) Analysed using unpaired t-test **=0.008. (G) Analysed using Wilcoxon t-test *=0.0312.

### Changes in the plasma polar metabolome during the acute and chronic phases of hookworm infection

We studied changes in the plasma polar metabolome of participants during the pre-infection (week 0), acute (week 8 – 12) and chronic phases (week 36) using untargeted high-resolution liquid chromatography-mass spectrometry (LC-MS). From the negative and positive electrospray ionization (ESI) modes, 40 and 66 MS (Fig. S6A) features were annotated respectively, based on mass spectral database matching with MS-DIAL’s built-in metabolomics databases (Data File S4). A total of 12 annotated metabolites including amino acids, amino acid metabolic byproducts and heme metabolism were detected (Fig. 6A). Thus, a total of 94 unique metabolites were annotated, broadly encompassing a diversity of chemical classes including: amino acids, nucleotides, amino acid metabolism products, organic acids, some dietary-related alkaloids, and exogenous drugs (e.g., paracetamol) (Data File S4). Investigation of the sample variation after correction for run-order using QC samples highlighted excellent quality of the data across both positive and negative ESI modes, as evidenced by tight clustering of pooled participant QC samples in the central area of the plot and well within the variance of the samples (Fig. S6B). Metabolic Set Enrichment Analysis (MSEA) was performed to identify and interpret patterns of metabolite concentration changes during the acute and chronic phases of hookworm infection, relative to the pre-infection phase. Acute phase of infection was significantly associated with the enrichment of metabolites relevant to galactose and tryptophan metabolisms (Fig. 6A; Table S4). Other metabolic pathways enriched include metabolism of amino acids (glutamine and glutamate; alanine and aspartate); glyoxylate & dicarboxylate compounds, although these associations were not significant (Table S4). In the chronic phase of infection, we observed a modest enrichment of selenocompound metabolism, biosynthetic pathway of amino acids, metabolism of amino acids, as well as the biosynthesis of ubiquinone and/or terpenoid-quinones (Fig. 6A; Table S5), however enrichment did not reach significance. To interpret the MSEA data in a biological-relevant manner, we performed complementary statistical methods after controlling for potential confounding covariates. We visualized variations amongst all the annotated polar metabolites over the course of infection using PCA (Fig. 6B). Clustering of the datasets were observed at the participant level and variation between participants appears greater than changes induced by hookworm infection. PERMANOVA analysis revealed that the participant gender (“Sex”) was the greatest source of variation in the data (PERMANOVA, p < 0.001) (Fig. 6B, right panel, Table S6), followed by inter-participants human variation (“Participant ID”) (PERMANOVA, p = 0.072) (Fig. 6B, Table S6), thus highlighting inherent covariates that need to be considered to correctly identify the main plasma metabolites that changed over hookworm infection. Accordingly, mixed-mode repeated measures two-way ANOVA analysis, controlling for “sex” and “Participant ID” (Table S7) subsequently revealed that out of the total of 94 annotated polar metabolites, the plasma levels of each of the following metabolites including kynurenine, melibiose, threonic acid, indoxyl sulfate and 5-methoxypsoralen, were significantly different between at least two of the timepoints during the course of hookworm infection. Kynurenine and melibiose are the major contributor to the top two metabolic pathways: galactose and tryptophan metabolisms; respectively (Fig. 6A), during the acute phase of infection. The levels of kynurenine (Fig. 6C, 6D), melibiose (Fig. 6C, Fig. S6C) and indoxyl sulfate (Fig. 6C & Fig. S6D) were increased during the acute phase, before decreasing to levels similar to those of baseline, during the chronic phase of infection. In contrast, levels of the xenobiotic, 5-methoxypsoralen, decreased over the course of hookworm infection (Fig. 6C, Fig. S6E). Threonic acid levels were elevated during the chronic phase of infection, relative to levels observed at baseline (Fig. 6C, Fig. S6F). We observed changes in plasma levels of kynurenine (Fig. 6F), and there was an overall trend of reduction in plasma tryptophan levels during the acute phase, although not reaching statistical significance (Fig. 6G). However, we observed an increase in Kynurenine/Tryptophan ratio, an indicator of increased activity of the rate-limiting enzyme responsible for converting tryptophan to kynurenine; indoleamine 2,3-dioxygenase (IDO), which shunts tryptophan down the kynurenine pathway (Fig. 6H). IDO activity is associated with immunosuppression and regulation. Immune cells can induce IDO activity for example through the expression of cytotoxic T lymphocyte antigen 4 (CTLA-4) that has been shown to be upregulated in helminth-infected population (*11*). Thus, we analyzed the expression level of CTLA-4 in different T cell subsets of PBMCs: wherein slight increases in CTLA-4 frequency in both CD25^+^ CD127-FoxP3^+^ T_REGS_ and CD4^+^ CCR4^+^ CXCR3^-^ T_H_2 cells positive for CTLA-4 at week 4 post-infection (Fig. 6I-J) were noticed; with no differences observed in frequencies of CCR6+ CCR4-CCXR3^-^ T_H_9 (Fig. 6K). Interestingly, T_REGS,_ T_H_2 and T_H_9 had increased expression (MFI) of CTLA-4 (Fig. 6L-N) in the acute phase of infection, wherein kynurenine/tryptophan ratio was elevated (Fig. 6H). In the T_H_2 cell subset (Fig. 6M) the increased expression of CTLA-4 is observed up to week 12 post-infection, with the increase being specific to the CRTH2^+^ subset (Fig. S6G) as no increase in CTLA-4 MFI was observed in the CRTH2^-^ subset (Fig. S6G).

**Figure 6:**
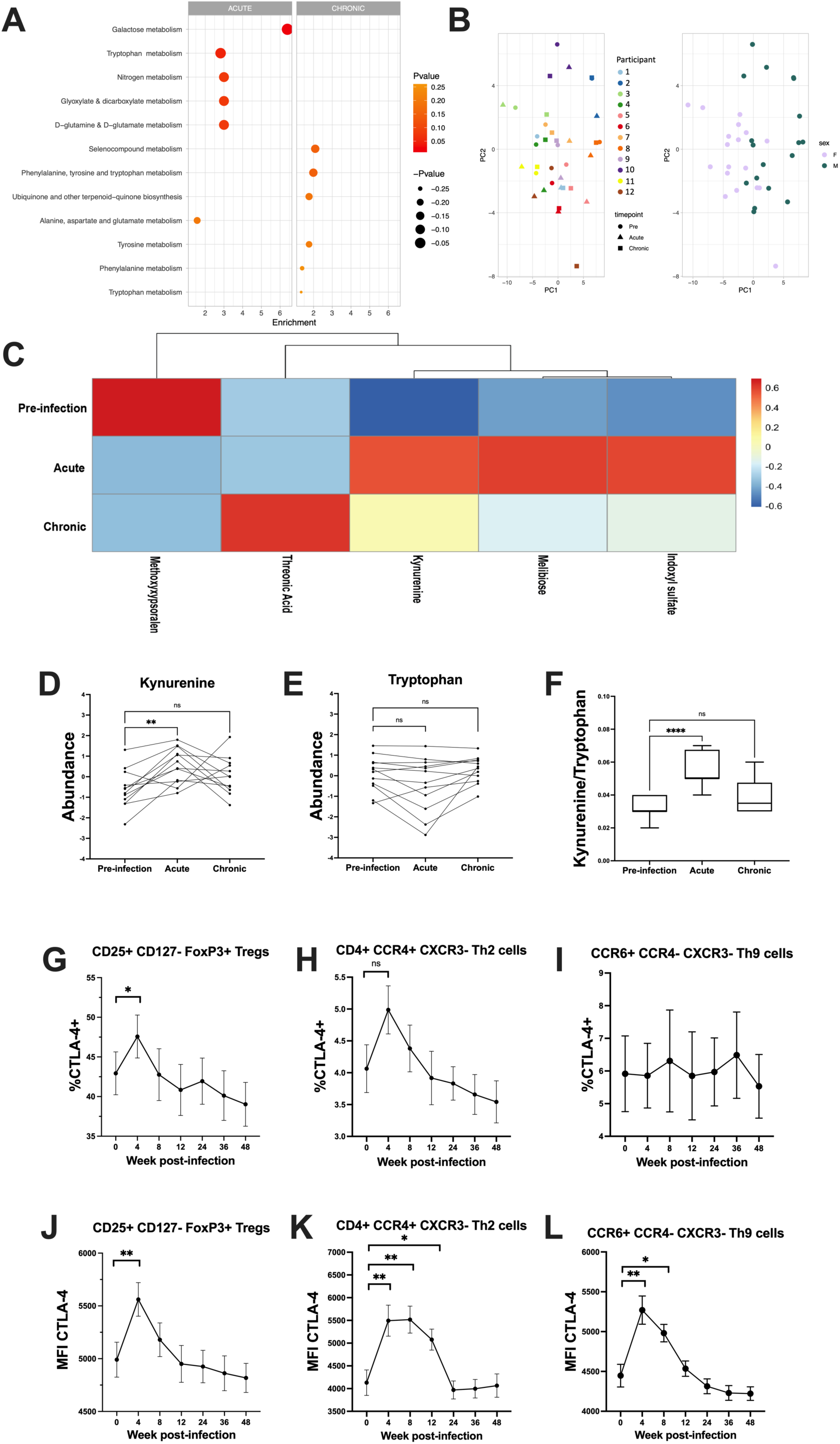
Changes in the plasma metabolome over the course of hookworm infection. (A) Top six enriched metabolic pathways identified using MSEA comparing pre-infection vs acute phase (left panel) or chronic phase of infection (right panel). (B) PCA scores plot on the plasma polar metabolome labelled by participant and by sex. (C) Heatmap of five annotated plasma metabolites significantly (p < 0.05) changed over the course of hookworm infection, as determined by mixed-mode, two-way analysis of variance (ANOVA), controlling for ‘Participant ID’ and ‘sex’ as covariates. (D) Post-hoc analysis indicating changes in plasma kynurenine levels over the course of hookworm infection. significantly different to pre-infection kynurenine levels by Dunnett’s post-hoc test. (E) Changes in plasma tryptophan levels over the course of hookworm infection. (F) Changes in plasma kynurenine/tryptophan ratio, a marker of indoleamine 2,3-dioxygenase (IDO) activity. Frequency of CTLA-4+ (G) CD25+ CD127-FoxP3+ T_REG_, (H) CD4+ CCR4+ CXCR3-T_H_2 cells and (I) CCR6+ CCR4-CXCR3-T_H_9 cells. (J-L) CTLA-4 MFI for population showed in G-H-I respectively. Graphs showing mean± SEM. In (D-F) values shown in the graph represent log_10_ transformed, auto scaled normalized data (N = 12 participants). Results were analysed with (F-G) Dunnett’s post-hoc test, (G-N) One-way ANOVA with Mixed-effect analysis with Tukey’s multiple comparison test. p= *<0.05, **<0.005, ***< 0.0005, ****<0.0001

## DISCUSSION

Since the first trial in 1984(*28*), experimental hookworm infections have been used in a range of healthy volunteer and human disease settings. To date, host peripheral blood immune responses have been studied due to limitations in obtaining specimens from tissues such as the gastro-intestinal tract. In addition, clinical intervention studies are difficult to develop due to the lack of sufficient knowledge of the immunological interplay between the infecting hookworm, the human host, and the disease context of the host. In this longitudinal study we examined healthy volunteers infected with *Na* for one year. We monitored symptoms and analyzed peripheral immune responses and microbiota. *Na* established a chronic infection in all participants, confirmed by eggs present in feces and adult worm visualization in the intestinal tract. Gastrointestinal symptoms appear to precede egg detection in the feces. Using cryopreservation methods (*25, 29*) we used reanimated cryopreserved larvae in four of our participants, we achieved a successful infection in all four participants and found no significant variation in immune responses. As cryopreservation did not alter infectivity, this method can be used to overcome some of the limitations when performing human controlled infection model such as long-term storage of infective larvae and preparation of defined batches for use in trials. This is the first study demonstrating that cryopreserved hookworm larvae can be used in a clinical study and cryopreservation has the potential to lend itself to cGMP manufacture and biobanking. Our studies show that *Na* infection induces a transient increase in peripheral blood eosinophils, with eosinophilia peaking at week 8 post-infection. Serum IL-5 levels also increase from week 4 post-infection, preceding the peak in blood eosinophils. Increased levels of the eosinophil degranulation products ECP and EDN were detected in feces and were associated with the increased levels of eosinophils. We used ECP and EDN as a proxy measure for eosinophilic intestinal inflammation, in combination with fCal as measure of neutrophilic inflammation. The timing of the increase in ECP and EDN found in fecal samples suggests that parasites start reaching the gut 4 weeks post infection, causing IL-5 dependent eosinophil responses whose inflammatory mediators are detectable in the feces. fCal increased after infection, but levels remained within the normal limits and did not correlate with neutrophils circulating in the peripheral blood. However, interestingly, calprotectin has been shown to be expressed by eosinophils in mice(*30*) and is elevated in eosinophilic gastrointestinal disorders(*31*) therefore the changes in fCal levels observed in the current study could be of eosinophilic origin. We also measured HNL in feces as a specific marker for neutrophil inflammation(*32*) and detected no increase in this marker, supporting the idea of a local, specific eosinophilic-driven response to hookworm infection. Measuring ECP, EDN, HNL and fCal in feces is a non-invasive method to detect local inflammatory responses during infection. We examined immune cells in the PBMC fraction and other cytokines in serum samples, and only detected serum IL-5 changes following *Na* infection. The transient nature of the increase in gut inflammation could possibly relate to the adult parasite in the gut being mature enough to start modulating the gut immune responses from week 12 of infection. For all parameters analyzed, such as cytokines, immune cells, microbiome, and metabolites, variation between individuals was greater than changes induced by the hookworm, reflecting the challenges involved in studying immune responses in humans(*33*). We found microbial alpha and beta-diversity to be stable during infection. Only 3 small studies have previously investigated microbial dynamics during a controlled *Na* infection. Our results are consistent with all previous studies, which have not identified a significant effect of infection on overall community composition or changes to alpha- diversity (*34–36*). In contrast to us, Ducarmon et al. identified a small increase in microbial richness alone (Chao1) from trial week 8-20, which our study did not replicate. Several studies have identified convincing microbiome signatures associated with natural hookworm infections (*37*). This seeming discrepancy is likely explained by a multitude of confounding factors such as hookworm burden, hookworm species, lifestyle, and hygiene, among others. Furthermore, the fecal microbiome may not capture the nuanced microbiome dynamics that may occur locally at the site of infection. A convincing understanding of the relationship between the hookworm and the microbiome will likely need larger sample sizes and small intestinal biopsies. Interestingly, we observed that moderate GI symptoms are associated with a more diverse microbiome, higher fCal is associated with increased diversity at the OTU level, and that changes in alpha-diversity are driven by OTUs belonging to the Firmicutes phylum. Similarly, in the longitudinal study by Ducarmon et al. GI symptoms were associated with modest differences in microbial diversity and microbiome stability(*34*). A deeper understanding of the functional diversity that differentiates between the responders and non-responders will allow us to better understand how the microbiome protects against hookworm-induced GI inflammation. Future studies using shot-gun metagenomics or RNA-sequencing will allow us to better understand microbiome functional potential in this context. We, then, used Smartpill^TM^ and observed a decrease in small bowel pH. The observed pH change could be associated with a change in mucus composition in the small intestine, as reported from mouse studies using *Trichuris muris,* where during the acute and chronic phases of infection different mucin expression is observed as well as glycoprotein hypersecretion driven by IL-13(*38*). However, pH was measured through Smartpill^TM^ which may not be sensitive enough or located correctly to measure changes in mucus pH. Nonetheless, the infection is well tolerated due to absence of adverse effects and no significant changes in intestinal physiology.

A novel feature of our study was the analysis of the levels of plasma polar metabolites using high-resolution LC-MS untargeted metabolomics methodology and determining whether they correlate with changes found in PBMC and cytokine immune parameters. MSEA analyses in plasma metabolites highlighted tryptophan metabolism as particularly enriched during the acute phase of infection, with kynurenine being the major contributor. We then considered that the increased plasma kynurenine was complemented partly through the conversion of tryptophan to kynurenine *via* the kynurenine pathway sub-branch of tryptophan metabolism (*39, 40*). Indeed, we observed changes in plasma kynurenine/tryptophan ratio, indicative of increased activity of the rate-limiting enzyme that converts tryptophan to kynurenine; indoleamine 2,3-dioxygenase (IDO), thought to be part of regulatory mechanisms to protect the host from an over-activated immune system(*41*). Tryptophan is a substrate for the generation of kynurenine, a metabolite best known for its association with inflammation, immune responses and neurotransmission(*39, 42*). Activation of the kynurenine pathway is associated with immunosuppression, reducing activity of NK cells, dendritic cells and T cells(*42*). While we observed an increase in kynurenine in the acute phase of infection, kynurenine biosynthesis is also associated with infection with influenza B and herpex simplex virus(*43*). Alteration in tryptophan metabolism has been observed in SARS-CoV2 positive subjects(*44*), thus changes in kynurenine biosynthesis could be a physiological change in response to infection rather than adult parasites inducing immunomodulatory pathway. Our findings pinpoint the need for future studies to conduct more in-depth, targeted metabolomic analyses quantifying changes in various aspects of tryptophan metabolism, particularly downstream metabolites of the kynurenine pathway, to better discern the potential role(s) of tryptophan metabolism products on the immunomodulatory potential of controlled hookworm infection as well as to distinguish the contrast the changes that are driven by a general response of the host to normal infections. In addition to enrichment of tryptophan metabolism, MSEA analysis also indicated enrichment of galactose metabolism during the acute phase of infection. This enrichment is exclusively driven by increased melibiose levels in plasma. Melibiose, a disaccharide of glucose and galactose, has been linked to suppression of T_H_2 cells and induction of oral tolerance in a model of OVA sensitization in mice(*45*) and may thus have involvement in reducing type 2 inflammation (e.g. IL-5 and eosinophils) during hookworm infection. In addition, immune-regulatory pathways have been associated with expansion of T_REGS_ and production of IL-10. In recent years, the surface protein CTLA-4 has been linked to immunomodulation. While we did not observe a general increase in T_REGS_ in peripheral blood, we detected an increase of CTLA-4+ T_REGS_ and T_H_2 cells 4 weeks post hookworm infection. Both T_REGS_ and T_H_2 cells have higher expression of CTLA-4 as measured by MFI, with T_H_2 having increased CTLA-4 expression up to 12-week post infection. Furthermore, we observed an increase in CTLA-4 expression on T_H_9 cells, but no increase in CTLA-4^+^ T_H_9 frequency. Decrease of these populations in the blood could reflect their migration into tissue like lungs and intestine, where *Na* transit through and reside, to reduce inflammatory responses and induce tissue repair. Increase in T_REGS_ has been observed in individuals infected with hookworm, and these cells expressed higher levels of CTLA-4, IL-10, and TGF-β(*46*). De Ruiter and colleagues did not observe expansion of T_REGS_ in helminth-infected Indonesian, however they showed expansion of CTLA-4+ T_REGS_ and deworming reduced the frequency of this population. Similarly, we observed expansion of CTLA-4+ T_REGS_, however in our study they decreased over the course of infection, without performing anti-helminth treatment. The main difference could be that multiple infection with different helminths since early age and re-infection over time, as normally happens in helminth endemic areas, drives a constant increase in this T_REG_ subpopulation. CTLA-4+ T_REGS_ have been shown to be suppressive of antigen-mediated lymphocytes expansion, indicating a potential role in parasite survival and immunomodulation(*46*), and possibly if we want to develop helminth as a therapeutic agent we should consider multiple re-infection to replenish the modulatory T_REGS_ pool. With this study we wanted to establish a controlled hookworm infection, with confirmation of *Na* infection and safety analysis as primary outcomes, and immune-profiling, microbiome, and metabolites as explorative outcomes. We proved safety and tolerability of infection in a healthy cohort in New Zealand, and the possibility with rigorous sampling to have a global picture of the effect of hookworm infection on the human host. A limitation in this study is that we are focusing on the systemic responses induced by *Na* at specific timepoints, therefore if cytokines are released outside the timepoint selected we will not be able to detect those changes. Similarly, if cytokines/chemokines are released locally in the intestinal tissue, these are unable to be detected. Nonetheless, this controlled infection is a useful tool to further study host-parasite interaction. The single *Na* inoculation used in this study provides a safe and robust method to study systemic and possibly local immune responses against hookworm and the use of helminths is being studied as a possible novel therapeutic agent (*47*). This controlled human hookworm model provides the base to bring forward hookworm therapies against inflammatory disorders and to build a model to test vaccine efficacy against *Na*.

## MATERIALS AND METHODS

### Study Design

The primary objective for this study was to establish a cohort of healthy individuals infected with 30 *Na* larvae and profile for 1 year at given time points, gut motility, immune cell, antibody, microbiome and metabolomic responses pre and post infection. This study was conducted in Wellington, New Zealand. This clinical study was approved by the Health and Disability Ethics Committee (Ethics reference 19/CEN/81) and registered on the Australian New Zealand Clinical Trials Registry (ACTRN12619001129178). Study outline and further information is found in Fig. 1 and Data File S1.

### Symptom questionnaire

Participant symptoms were monitored by symptom responses using a modified version of the Talley gut symptom questionnaire(*48*) on weeks 1-12. Symptoms were graded as Mild (Nagging or annoying), Moderate (strong negative influence on daily living) and Severe (disabling) and were self-reported by each participant. In addition, participants were asked to photograph skin reactions during the first 4 weeks of infection.

### Parasitology

Two methods were employed for producing *Na* infective larvae. Fresh larvae were produced using fecal-charcoal cultures. Feces collected from a chronically infected volunteer who was screened for communicable agents, were mixed with activated charcoal, and incubated for 7-10 days at 26°C. For “Cryoworms”, reanimated cryopreserved larvae were developed to infective larvae using methods previously described(*25, 29*). Recovered larvae were isolated and washed in 5% iodine solution followed by several washes with sterile distilled water. A dose of 30 *Na* infective larvae (iL3) was used for each participant, administered through two dressings each containing 15 iL3 applied on each forearm. Detection of patency was carried out through presence of *Na* eggs in the feces of infected individuals using the McMaster technique from week 6 following application of larvae.

### Whole blood

Whole blood immunophenotyping was performed using 600 µL of heparinized whole blood. RBCs were lysed using in-house made ACK Lysis Solution (ddH_2_O with 1mM EDTA, 150mM NH_4_Cl, 10mM NaHCO_3_ at pH 7.4) at 4°C for 10 minutes. Samples were then diluted in PBS to arrest cell lysis, followed by centrifugation at 500g for 5 min at 4°C. The RBC lysis step was repeated once if necessary. Cells were washed twice with PBS and counted. Cells were stained with 1:2000 Zombie NIR Fixable Viability dye (Biolegend) in the dark at room temperature for 15 minutes. Subsequently, cells were stained with the antibody mix (Data file S2) in FACS buffer (PBS + 2% FBS, 0.01% NaN₃) in the presence of 20% Brilliant Stain Buffer Plus (BD Biosciences), at 4°C for 20 minutes. Prior to staining, the antibody mix was filtered through a 0.1µm Centrifugal Filter (Sigma). Subsequently, cells were washed twice with FACS buffer and analyzed on a 5-laser Aurora spectral flow cytometer (Cytek Biosciences). Data was analyzed using FlowJo v10.7.1 and Prism V8 (GraphPad Software Inc.).

### PBMC isolations and analysis

Peripheral blood mononuclear cells (PBMCs) were isolated from heparinized whole blood by SepMate™ PBMC Isolation Tube (STEMCELL Technologies). Briefly, whole blood was diluted 1:1 in PBS +2% FBS, added to the SepMate™ PBMC Isolation Tube containing 15ml of Lymphoprep (STEMCELL Technologies) and centrifuged at 1200g for 12 minutes at RT. Collected PBMC were washed three times with PBS + 2% FBS and cryopreserved in FBS + 10% DMSO. Cryovials containing PBMC suspension were transferred to a Corning^®^ CoolCell^®^ Container, which was placed at −80°C overnight. Cryovials were then stored in liquid nitrogen until analysis. On day of analysis, thawed cells were resuspended in PBS + 2% FBS, counted and for optimal protocol up to 5×10^6^ cells were resuspended in FACS buffer and used for staining. Two antibody panels were designed to assess 1) T cell subsets, NK T cells, MAIT cells and 2) B cells, NK cells, Monocytes and Dendritic cells. Details on antibodies used and gating strategies are listed in Data File S2 and Figure S3. PBMCs were incubated with 1:2000 Zombie NIR Fixable Viability dye (Biolegend) at room temperature for 20 minutes. After washing, cells were incubated with 1:40 FC block (Biolegend) for 10 minutes at room temperature.

For panel 1, cells were surface stained for CD1d and MR1 tetramers for 15 minutes at 37°C, washed and sequentially stained for CD25, TCRγδ, CXCR3, CCR10, CCR6, CCR4, CCR7 for 5 minutes each at 37°C. For panel 2, cells were sequentially stained for CXCR5 and CCR7 for 5 minutes each at 37°C. After sequential staining cells were incubated with surface receptor mix for 20 minutes at room temperature in the presence of Brilliant Buffer Plus (BD Biosciences) and Monocyte Block (1:30, Biolegend) only for panel 2. Cells were fixed and permeabilized with FoxP3/Transcription Factor Staining Buffer Set (eBioscience) for 30 minutes at room temperature and subsequently incubated with FC block (1:40, Biolegend) for 10 minutes at room temperature. Cells were washed in Permeabilization Buffer (eBioscience), incubated with intracellular antibody mix for 40 minutes on a shaking plate at room temperature. Cells were then resuspended in FACS buffer and analyzed on a 5-laser Aurora spectral flow cytometer (Cytek Biosciences).

### PillCam

Hookworm visualization in the small bowel was carried out using the PillCam^TM^ SB3 Capsule Endoscopy System (Medtronic). Capsules were administered by supervision of trained staff from The Rutherford Clinic (Lower Hutt, New Zealand). Participants were instructed to drink a bowel prep (Klean-Prep), maintain a clear liquid diet from midday prior to administration and fast from midnight prior to administration. PillCam^TM^ video recordings were analyzed using RAPID Reader software by two people separately, then final numbers of worms visualized were discussed and finalized. Data were tabulated and graphically presented using Prism V8 (GraphPad Software Inc.).

### Smartpill delivery

Motility, temperature, pH and pressure of gastrointestinal tracts were measured at indicated timepoints using the SmartPill^TM^ Motility Monitoring Capsule and SmartPill^TM^ Software v3.1 (Given Imaging and Medtronic). Capsules were administered by trained staff according to the User Manual (DOC-4092-01 November 2017) and software instructions. Participants were instructed to fast from food and drink for 8 hours before a test, ate a SmartBar^TM^ prior to capsule ingestion and were instructed to refrain from eating until 6 hours into the test. Tests were analyzed using the software’s Test Analysis Wizard.

### Microbiome

Fecal samples were self-collected using a disposable toilet accessory (DNA Genotek, Canada) and samples were placed into OMNIgene.GUT tubes (DNA Genotek, Canada) according to manufacturer’s instructions. Samples were placed in chiller-bags containing an ice-pack and delivered to the laboratory within 24 hours of collection. OMNIgene.GUT tubes were stored at –80°C until processing. DNA was extracted from fecal samples using the Qiagen MagAttract PowerSoil DNA KF Kit optimized for the Thermofisher KingFisher robot. DNA quantification and quality checks were done via Qubit. 16s rDNA marker gene (V4) was amplified using the high-fidelity Phusion polymerase using dual-barcoded primers(*49*) and PCR products were verified by gel electrophoresis. PCR reactions were cleaned and normalized using the SequalPrep 96-well Plate Kit. Library samples were pooled and sequenced on the Illumina Miseq. Fastq files were quality filtered and clustered into 97% similarity operational taxonomic units (OTUs) using the mothur software package(*49*). Representative OTU sequences were assigned taxonomic classification using a Bayesian classifier against the Ribosomal database project (RDP) database(*50*). Before filtering, alpha diversity (Shannon measure) was calculated and plotted using Phyloseq(*51*). Data was centered log-ratio (CLR) transformed and filtered. Data was further analyzed in R using custom scripts and plotted using ggplot2(*52*).

### Plasma Metabolomics analysis

Plasma samples for metabolomics analysis were sent to AgResearch (Palmerston North, NZ). For acute phase of infection, week 8 samples were analyzed except for 2 participants for which week 12 samples were analyzed due to inability of collecting week 8 samples due to COVID-19 restrictions in New Zealand. For chronic phase week 36 samples were analyzed. Polar metabolites were extracted from participant plasma samples as previously described (*53*). Briefly, processed plasma was placed into an HPLC vial for LC-MS using HILIC chromatography. Pooled quality control (QC) samples were prepared by pooling together 50 µL of each participant plasma sample. QC were analyzed alongside samples to allow for signal normalization to correct for any loss of signal across the analytical batch.

Polar metabolites extracted from plasma samples were analyzed using a Shimadzu LCMS-9030 mass spectrometer equipped with a Shimadzu Nexera-x2 UHPLC system. Data were captured, converted and peaks were then annotated using the DIA MS/MS data collected against the built-in human metabolite library containing 13,303 MS/MS spectra(*54*). The resultant peak intensity table underwent run-order correction and normalization using pooled QC samples and the locally weighted scatterplot smoother (LOWESS) regression model. Plasma metabolome data (Data File S4) underwent log-transformation and autoscaling prior to the following analyses: (i) Metabolite Set Enrichment Analysis (MSEA) using the Quantitative Enrichment Analysis (QEA) module on MetaboAnalyst 5.0 (*55*); (ii) principal component analysis (PCA); (iii) permutational multivariate analysis of variance (PERMANOVA); (iv) mixed-mode repeated measures two-way ANOVA with Dunnett’s post-hoc multiple comparison tests, all analyzed using R software and/or GraphPad Prism (Version 8).

### ELISA for fecal degranulation products

For extraction of ECP, EDN and HNL from feces the granule protein extraction protocol was used as described elsewhere (*56*). Feces samples were self-collected and immediately frozen at −80°C. On the day of processing feces samples were thawed overnight in the refrigerator or at room temperature for 1 hour. Feces was weighted and diluted five times in extraction buffer consisting of phosphate-buffered saline, pH 7.4, supplemented with 10-mmol/L ethylenediamin-etretraacetic acid, 0.2% *N*-cetyl-*N,N,N*-trimethylammonium bromide (CTAB)(Sigma), 20% glycerol (Sigma), 0.05% Tween20, and 1% bovine serum albumin. The mixture was homogenized using a homogenizer mixer until a homogenous solution was obtained. After incubation at 4°C for 30-45 min and mixing, the homogenate was centrifuged at 4,000g for 30 min at 5°C. The supernatant was aliquoted and frozen at −20°C for later ELISA analysis. For each sample a separate tube of the diluted homogenate was weighed and centrifuged. By weighing the pellet obtained after discarding the supernatant, a measure of semidry weight was obtained. Fecal ECP, EDN and HNL were measured by ELISA (Diagnostics Development AB, Uppsala, Sweden) according to the manufacturer’s protocol. The levels of markers in feces were adjusted for water content and expressed as micrograms per gram of semidry faeces. For extraction of fecal calprotectin CALEX Cap devices (Bühlmann Laboratories, USA) were used according to the manufacturer’s protocol. Fecal calprotectin was measured by fCal ELISA (Bühlmann Laboratories, USA) according to the manufacturer’s protocol.

### ELISA/Multiplex Assays (Serologic analysis)

Plasma was separated from heparinized whole blood by centrifugation at 2000g for 15 minutes at 4°C. Serum was separated from clotted blood by centrifugation at 2000g for 10 minutes at 4°C. Plasma and serum were stored at −80°C until analysis.

For serological analysis, samples were analyzed using MILLIPLEX® MAP Kits purchased from Merck-Millipore. Serum samples were tested for IgE, IgG isotypes, IgA, IgM, RANTES, BCA-1, Eotaxin-3, TSLP, IL-33, Eotaxin, IFNγ, IL-1β, IL-3, IL-4, IL-5, IL-6, IL-8, IL-10, IL-12.p70, IL-13, IL-17A, TNFα, and TGF-β, according to manufacturer instruction. Briefly, 25ul of diluent containing antibody-coupled beads was added to a black clear-bottomed 96-well plate (Merck-Millipore). Diluted serum was added to the plate and incubated for 1 or 2 hours according to the kit protocol. Plates were washed, incubated with detection antibody, followed by another wash and incubation with Streptavidin-Phycoerythrin (Merck-Millipore). The plate was placed on a magnetic plate holder (Bio-Rad) for 1 minute before 35 µL of fluid was removed from wells and 150 µL sheath fluid (Thermo Fisher Scientific) added. Finally, the plate was analyzed in a Luminex 200 array reader (Merck-Millipore), and results were determined against commercial standards (Merck-Millipore) using the Luminex xPONENT® software. Cytokines under the limit of detection not shown in the result section.

For *Na*-specific IgG ELISA, samples were analyzed following the protocol previously described(*20*). Briefly, somatic extract was prepared using approximately 75000 iL_3_ in 500 µl lysis buffer (3 M urea, 0.2% SDS, 1% Triton X-100, 50 mM Tris-HCl). Larvae were homogenized by hand using a tissue grinder and subsequently sonicated at 4 °C. Sonicated larvae were centrifuged at 10000 g at 4 °C for 10 minutes, supernatants collected, and protein quantified by BCA assay (Pierce). 96-well Nunc MaxiSorp ELISA plates (Thermo Fisher Scientific) were coated with 5 µg/mL somatic extract overnight at 4°C. Plates were washed with PBST/0.05% BSA and blocked with PBST/5% BSA for 2 hours at room temperature with shaking (400 revolutions per minute/rpm). Plates were washed as above and incubated with 50 µL participant sera diluted at 1:100 in PBST/1% BSA for 1 hour at room temperature with shaking (400 rpm). Plates were washed as above and incubated with 50 µL goat anti-human IgG-horseradish peroxidase (Sigma-Aldrich) diluted at 1:5000 in PBST/1% BSA for 1 hour at room temperature with shaking. Plates were then washed and incubated with TMB substrate (Thermo Fisher Scientific) for 5 minutes. The reaction was stopped with an equal volume of 1M H_2_SO_4_ and absorbance was measured at 450 nm through Infinite® M1000 plate reader using i-control™ 2.0 software (Tecan).

### Analysis, visualization, and Statistical analysis

Whole blood flow cytometry data was analyzed using FlowJo software. PBMCs flow cytometry data was batch-to-batch normalized using CytoNorm algorithm in R studio. Data was analyzed using flowSOM clustering algorithm and UMAP algorithm as a visualization tool using the cloud-based OMIQ software (San Francisco, USA). Heatmaps and PCA plots were generated using R software or OMIQ and modified in Adobe Illustrator. Statistical tests were performed in R or using Graphpad Prism version 9 as indicated in each figure legends. D’Agostino & Pearson normality test were performed for each dataset before running One-way ANOVA.

## Data Availability

All data produced in the present study are available upon reasonable request to the authors

## Supplementary Materials

**Figure S1:** Skin reaction, egg production and worm count

**Figure S2:** Eosinophils correlation, cytokines, and immunoglobulins profiling

**Figure S3:** PBMCs gating strategies

**Figure S4:** Immuno-profiling of PBMC subpopulations

**Figure S5:** Microbial diversity and gut physiology

**Figure S6:** Changes in plasma levels of four polar metabolites and changes in CTLA-4 MFI of CRTH2+ or CRTH2-CTLA-4+ Th2 cells over the course of hookworm infection

**Data file S1:** Participant Inclusion/exclusion criteria

**Data file S2:** List of flow cytometry antibodies

**Data file S3:** OTUs data file.

**Data file S4**: Accurate masses and retention times of all annotated metabolites detected in plasma as analyzed using negative and positive electrospray ionization modes.

Data file S5: Primary data.

**Table S1:** Demographic Information

**Table S2:** Microbiome Dataset

**Table S3:** Microbiome Dataset Phylum and Symptoms score

**Table S4.** Metabolic Set Enrichment Analysis (MSEA) statistics comparing metabolic pathways enriched during the “acute phase” of hookworm infection, relative to baseline (i.e., “pre-infection phase”)

**Table S5:** Metabolic Set Enrichment Analysis (MSEA) statistics comparing metabolic pathways enriched during the “chronic phase” of hookworm infection, relative to baseline (i.e. “pre-infection phase”)

**Table S6:** PERMANOVA Analysis identifying “Sex” and “Participant ID” as important covariates to control for in downstream statistical analyses

**Table S7:** Two-way, repeated-measures Mixed ANOVA analysis statistics, controlling for “sex” and “participant ID” as covariates.

**Movie S1**: PillCam^TM^ video from hookworm-infected volunteer

## Acknowledgments

We would like to acknowledge the Hugh Green Cytometry Centre for their help with the cytometry data and Dr. Karl Fraser (AgResearch Ltd, Palmerston North, New Zealand) for providing help and insightful discussion in plasma metabolomics analyses. We would also like to acknowledge Prof. Alex Loukas and Dr. Paul Giacomin for insightful discussion.

## Author contributions

Conceptualization: MC, GLG, SI

Funding Acquisition: GLG

Formal Analysis: FV, BL, SLN, AC, LFF, JST, TCM, MC

Investigation: FV, BL, SLN, KM, BY, TTK, JM

Writing – Original Draft: FV

Writing – review & editing: FV, BL, SLN, TCM, LFF, AC, JST, OG, GLG, MC, SI

Project administration: MC

## Data and materials availability

All data are available in the main text or the supplementary materials.

---

**Supplementary Figure 1:**
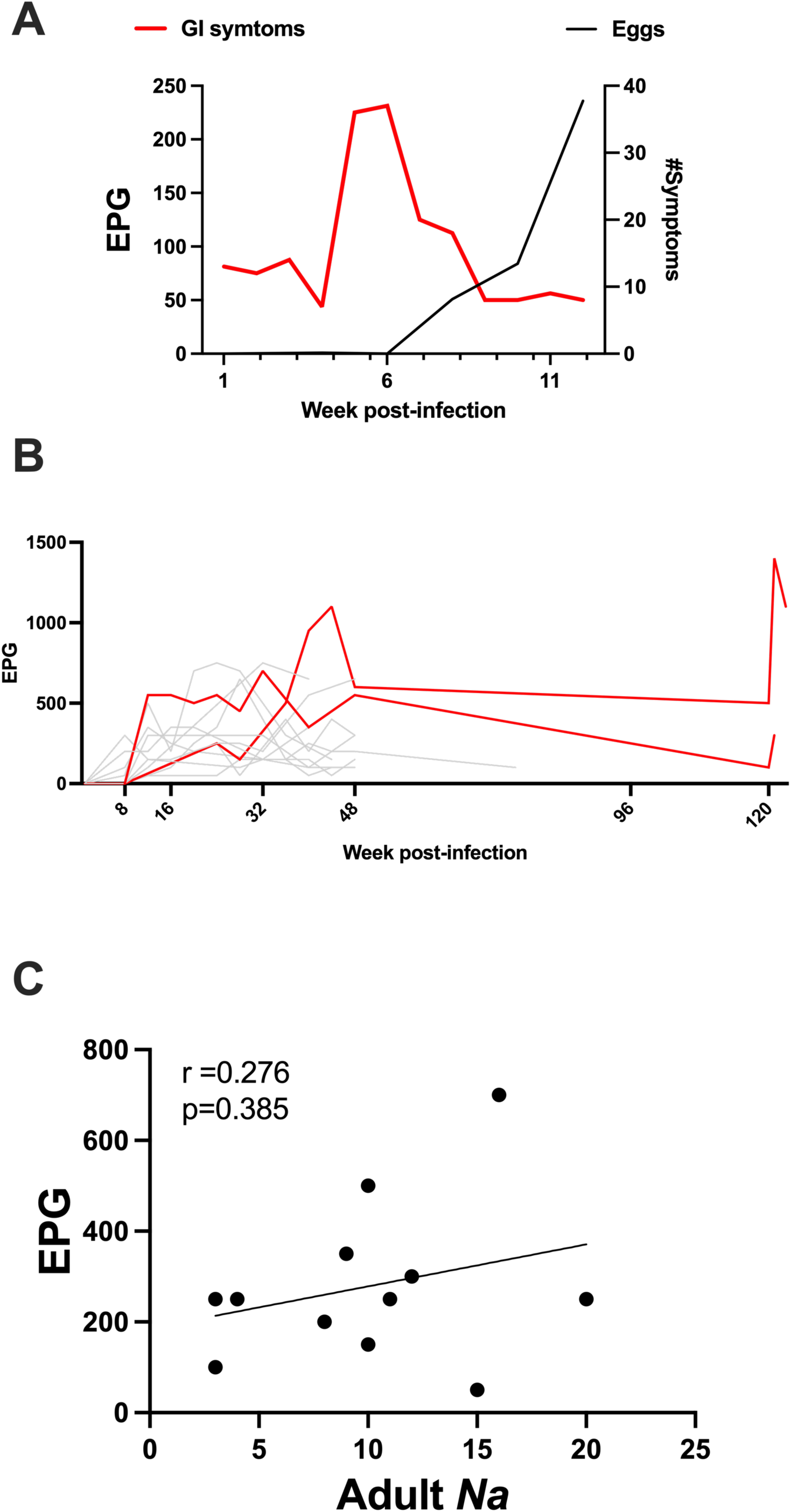
Skin reaction, egg counts and adult worm enumeration. (A) GI symptoms (shown in Fig. 2B) plotted against mean egg count. (B) Egg counts highlighting 2 participants (red lines) with patent infection over 120-week post-infection. (C) Correlation between EPG and adult *N. americanus* visualised in the intestine.

**Supplementary Figure 2:**
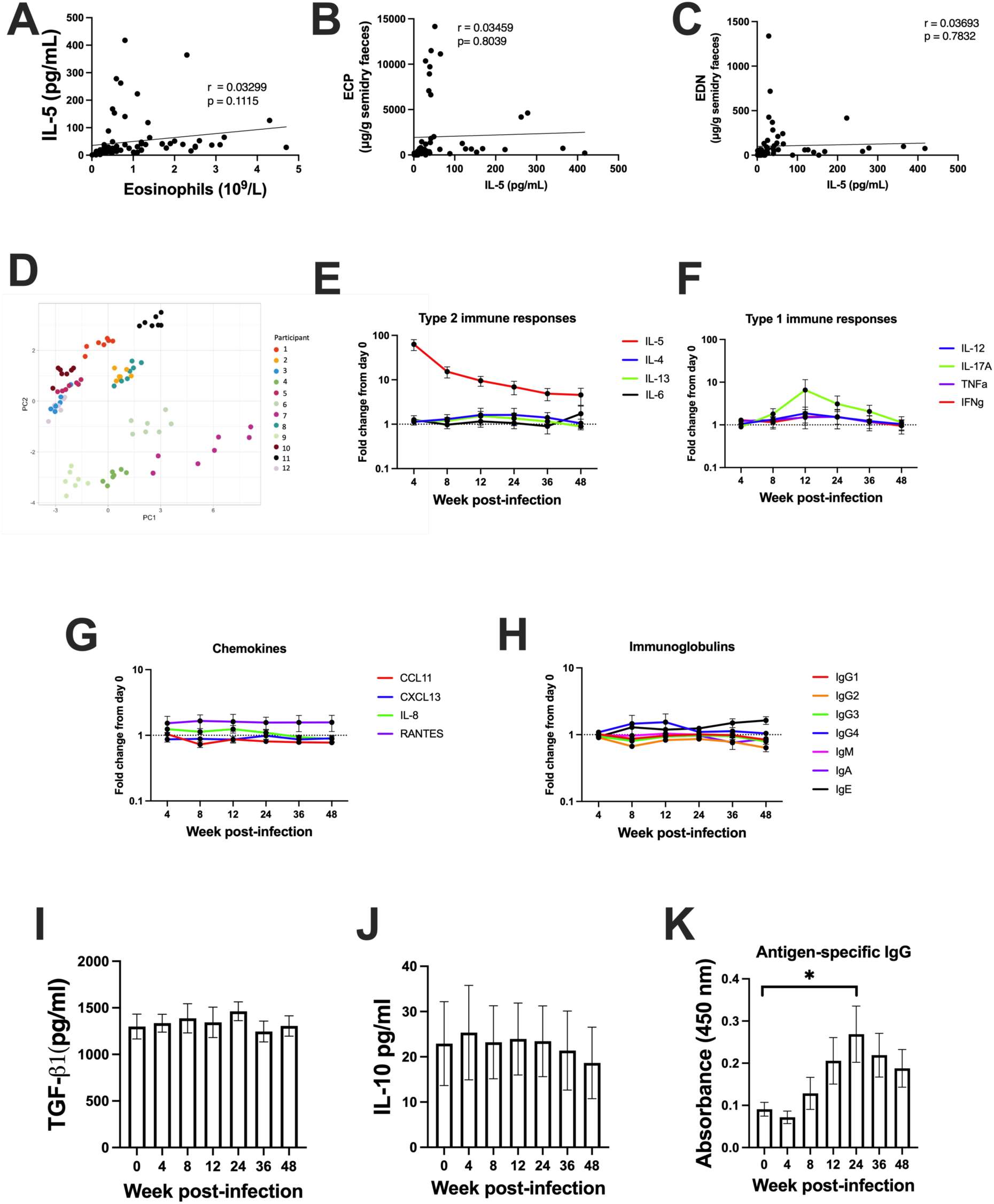
Cytokine panels during hookworm infection. IL-5 correlation with (A) eosinophils, (B) ECP, (C) EDN during Na infection. (D) PCA plot created with R software to visualise Luminex data for cytokines and chemokines analysed during the trial. (E-H) Graph showing fold changes from baseline (day 0 – uninfected) of type 2 cytokines (E), type 1 cytokines (F), chemokines (G) and antibodies (H) over the course of infection. (I) Graph showing mean ± SEM of TGF-β1 measured by Luminex (N=8). (J) Mean ± SEM of IL-10 measured in serum by Luminex (N=7). (K) Bar graph showing mean ± SEM of antigen-specific IgG, measured by antigen-specific ELISA.

**Supplementary Figure 3:**
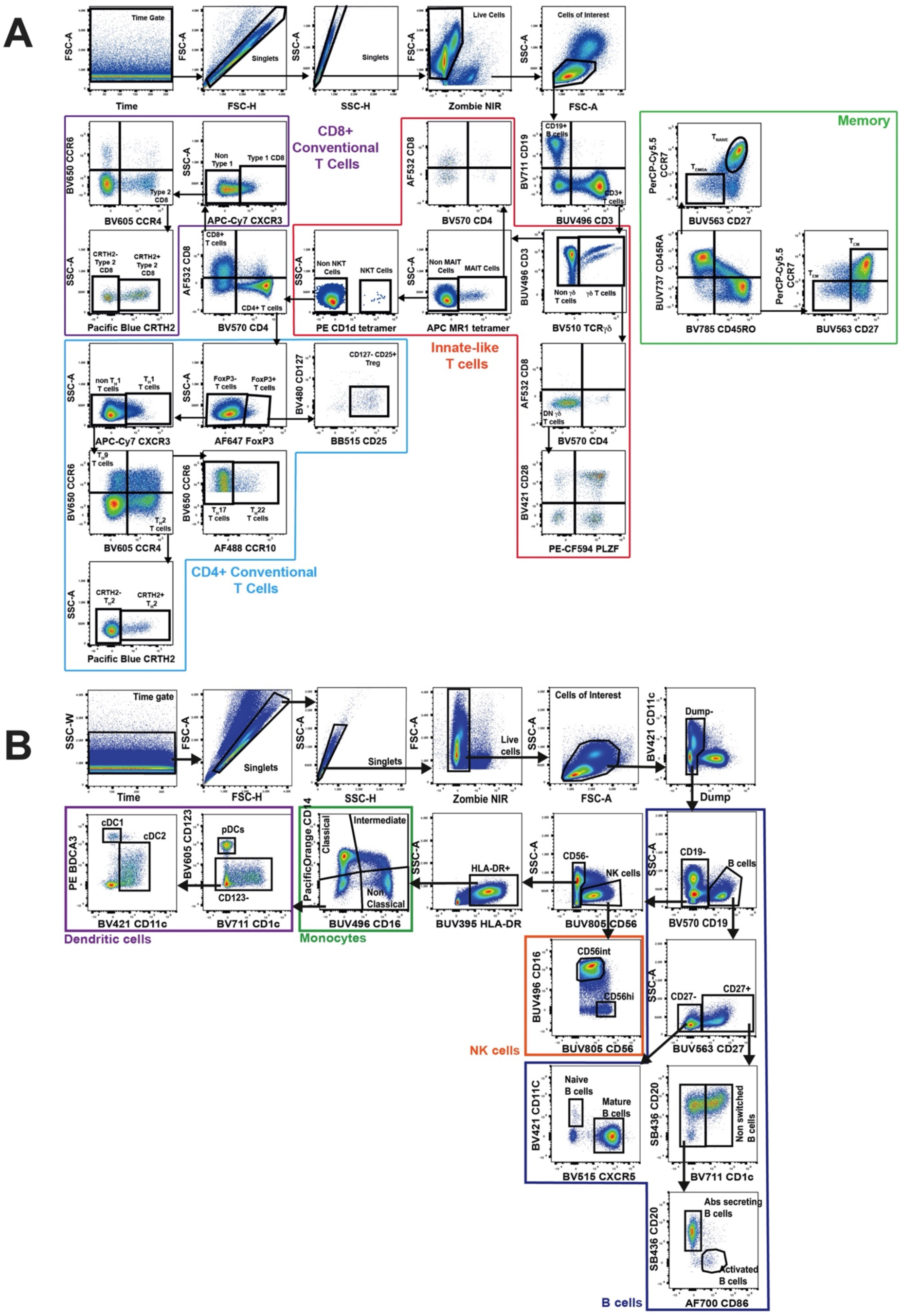
PBMCs gating strategies. (A) Gating strategies for PBMCs panel 1. (B) Gating strategies for PBMCs panel 2.

**Supplementary Figure 4:**
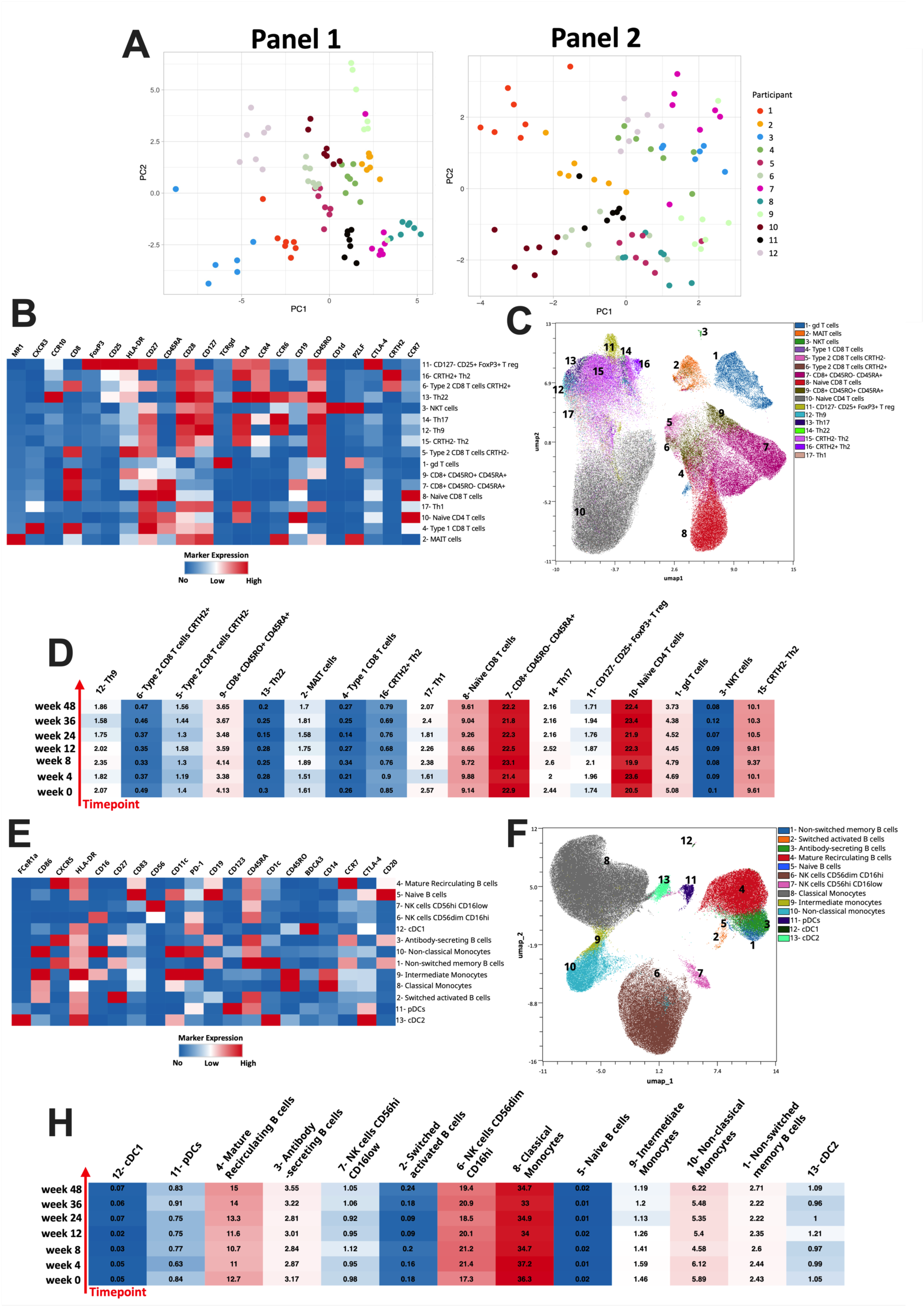
PBMC subpopulations unchanged during hookworm infection. (A) Immune cells variability visualized by PCA plot generated using R software. (B) Heat map summary of median expression values of cell markers expressed by subpopulations identified with panel 1. (C) UMAP generated using OMIQ with overlapped panel 1 subpopulations. (D) Frequencies of subpopulations per timepoints. (E) Heat map summary of median expression values of cell markers expressed by subpopulations identified with panel 2. (F) UMAP generated using OMIQ with overlapped panel 2 subpopulations. (H) Frequencies of subpopulations per timepoints. In (D) and (H) color gradient represent the percentage of population, with red showing the highest and blue the lowest.

**Supplementary Figure 5:**
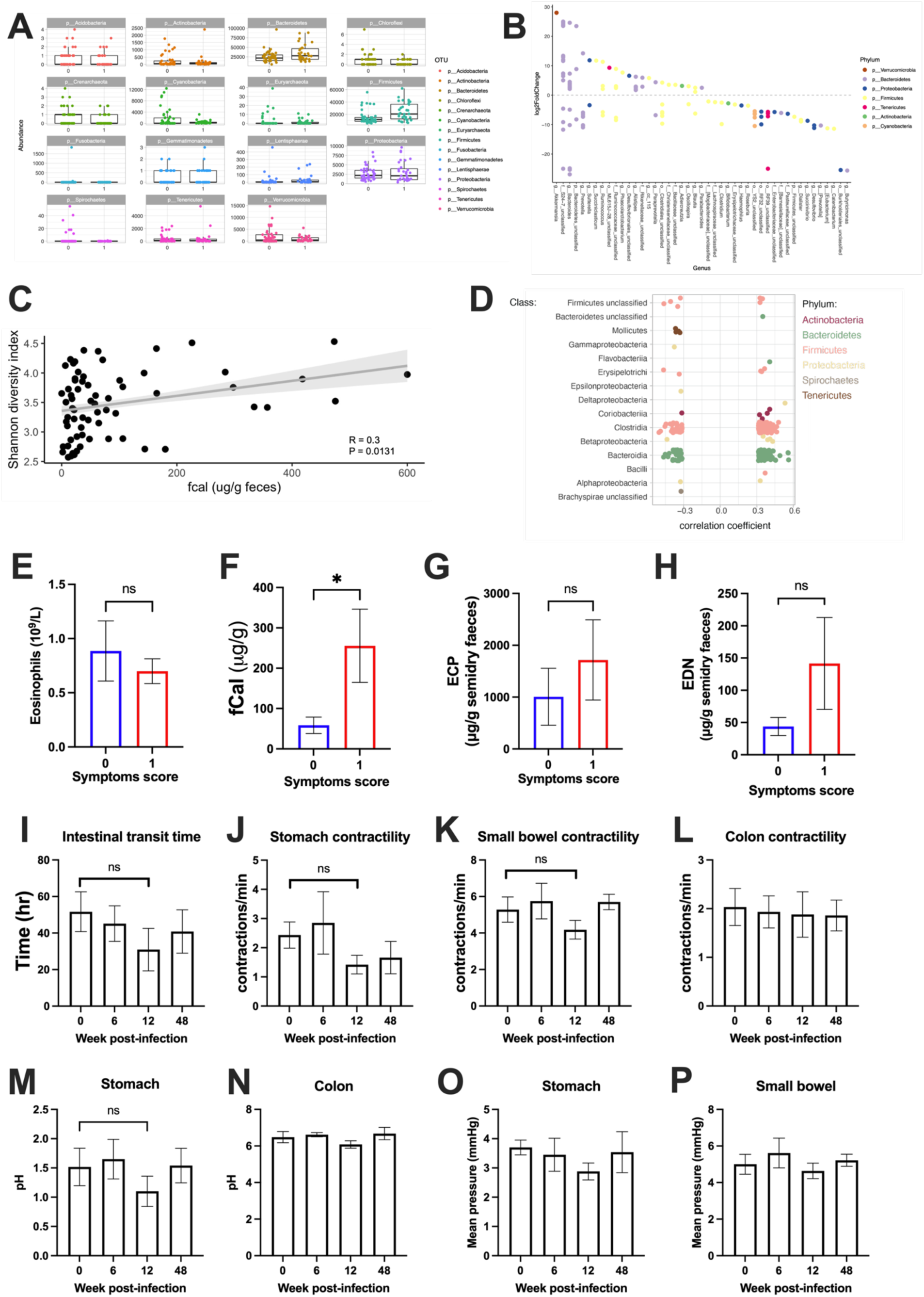
Changes in microbiome are associated with moderate symptomology after hookworm infection without changes in gut physiology. (A) Relative abundance of the top 100 most abundant taxa split by those who experience mild to moderate symptoms (symptom score 0) and those who experience severe symptoms (symptom score 1) and faceted by Phylum. (B) OTUs that were identified to be significantly different between those who experience mild to moderate symptoms (symptom score 0) and those who experience severe symptoms (symptom score 1) as determined using DESeq2 with an adjusted P value < 0.01. (C) Spearman correlation between an individual’s microbiome diversity (Shannon index) and fCal levels. (D) OTUs that were identified to have a significant Spearman correlation (P < 0.01) with fecal calprotectin levels. Correlation coefficient is plotted and OTUs are arranged by Class. Color represents Phylum. (E-H) Mean ± SEM of eosinophils count (E), fCal (F), ECP (G) and EDN (H) according to symptoms score. (I-P) Gut physiology measured through Smartpill^TM^ (N=6), bar graph showing mean ± SEM for intestinal transit time (I), stomach contractility (J), small bowel contractility (K) and colon contractility (L). Changes in pH in the stomach (M) and colon (N). Mean pressure in the stomach (O) and small bowel (P). (E-H) Analysed using Mann-Whitney t-test, (I-P) analysed using one-way ANOVA with Kruskal-Wallis multiple comparison test. *<0.05

**Supplementary Figure 6:**
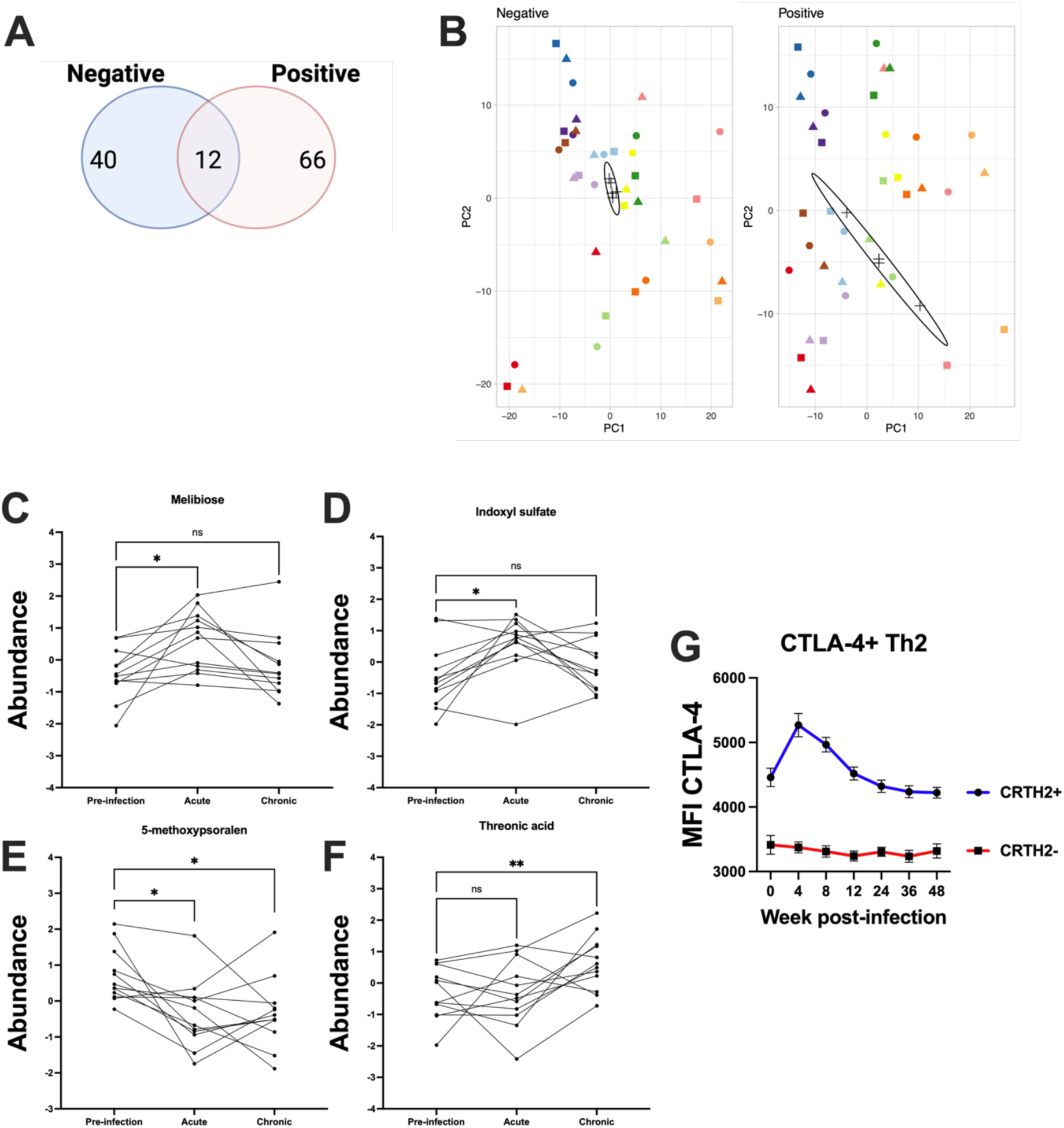
Changes in polar metabolites and CTLA-4 expression. (A) Number of annotated plasma metabolites detected in the negative or positive electrospray ionization (ESI) modes, including those detected in both modes. (B) Data overview and quality assurance demonstrating good method reproducibility in the negative (left panel) and positive ESI (right panel) modes, as reflected by tight clustering of QCs evident in the principal component analysis (PCA) scores plots. Changes in plasma levels of (C) Melibiose, (D) indoxyl sulphate, (E) 6-Methoxypsoralen, or (F) threonic acid, over the course of hookworm infection.*p< 0.05; **p < 0.01, significantly different to pre-infection kynurenine levels by Dunnett’s post-hoc test, following Mixed ANOVA analysis. Values shown represent log_10_ transformed, auto scaled normalized data (N = 12 participants). (G) Graph showing expression of CTLA-4 (MFI) for CRTH2+ (blue) and CRTH2-(red) CD4+ CCR4+ CXCR3-T_H_2 cells. Values shown are mean± SEM (N = 12 participants).

**Table S1:** Demographic Information.

|  |  |
| --- | --- |
| ***n*** | 13 |
| **Age (years): Median [Range]** | 47 [19-64] |
| **Gender** |  |
| **Male** | 6 (46.2) |
| **Female** | 7 (53.8) |

**Table S3:** Microbiome Dataset Phylum and Symptoms score.

|  | Phylum | mild symptoms | moderate symptoms | statistic | df | p | p.adj | p.adj.signif |
| --- | --- | --- | --- | --- | --- | --- | --- | --- |
| 1 | p_Crenarchaeota | 0 | 1 | 2.65023609 | 74.9007931 | 0.00981 | 0.04905 | * |
| 2 | p_Cyanobacteria | 0 | 1 | 2.78380459 | 48.4518919 | 0.00764 | 0.04905 | * |
| 3 | p_Firmicutes | 0 | 1 | -3.0583608 | 55.3237504 | 0.00343 | 0.04905 | * |
| 4 | p_Bacteroidetes | 0 | 1 | -2.339609 | 51.1879021 | 0.0232 | 0.065 | ns |
| 5 | p_Chloroflexi | 0 | 1 | 2.27891505 | 63.6827473 | 0.026 | 0.065 | ns |
| 6 | p_Spirochaetes | 0 | 1 | 2.42550735 | 44.0284889 | 0.0195 | 0.065 | ns |
| 7 | p_Verrucomicrobia | 0 | 1 | 1.99772006 | 60.1674897 | 0.0503 | 0.10778571 | ns |
| 8 | p_Acidobacteria | 0 | 1 | 0.93065836 | 73.2509802 | 0.355 | 0.52090909 | ns |
| 9 | p_Actinobacteria | 0 | 1 | 0.88037904 | 66.5237391 | 0.382 | 0.52090909 | ns |
| 10 | p_Euryarchaeota | 0 | 1 | 1.0100773 | 61.7175791 | 0.316 | 0.52090909 | ns |
| 11 | p_Fusobacteria | 0 | 1 | 1.03470669 | 44.0000997 | 0.306 | 0.52090909 | ns |
| 12 | p_Lentisphaerae | 0 | 1 | -0.802856 | 71.5791369 | 0.425 | 0.53125 | ns |
| 13 | p_Gemmatimonadetes | 0 | 1 | -0.5939425 | 57.3842922 | 0.555 | 0.59464286 | ns |
| 14 | p_Proteobacteria | 0 | 1 | -0.6453448 | 50.9799637 | 0.522 | 0.59464286 | ns |
| 15 | p_Tenericutes | 0 | 1 | -0.1917242 | 46.5644433 | 0.849 | 0.849 | ns |

**Table S4.** Metabolic Set Enrichment Analysis (MSEA) statistics comparing metabolic pathways enriched during the “acute phase” of hookworm infection, relative to baseline (i.e. “pre-infection phase”)

| No | Metabolic pathway | Total compounds<sup>a</sup> | Hits | Q-statistic | Expected Q-Statistic | Enrichment Ratio<sup>b</sup> | Nominal P-value | Holm P-value | FDR<sup>c</sup> |
| --- | --- | --- | --- | --- | --- | --- | --- | --- | --- |
| 1 | Galactose metabolism | 27 | 1 | 27.77 | 4.35 | 6.388 | 0.0081 | 0.285 | 0.285 |
| 2 | Tryptophan metabolism | 41 | 3 | 12.26 | 4.35 | 2.821 | 0.0496 | 1 | 0.581 |
| 3 | D-glutamine & D-glutamate metabolism | 6 | 1 | 13.04 | 4.35 | 2.999 | 0.083 | 1 | 0.581 |
| 4 | Glyoxylate & dicarboxylate metabolism | 32 | 1 | 13.04 | 4.35 | 2.999 | 0.083 | 1 | 0.581 |
| 5 | Nitrogen metabolism | 6 | 3 | 13.04 | 4.35 | 2.999 | 0.083 | 1 | 0.581 |
| 6 | Alanine, aspartate and glutamate metabolism | 28 | 2 | 6.84 | 4.35 | 1.574 | 0.196 | 1 | 0.809 |

<sup>a</sup>Total number of metabolites deposited in the KEGG metabolic pathway database
<sup>b</sup> **Enrichment ratio:** Ratiometric enrichment expressed as acute phase/pre-infection phase
<sup>c</sup> P-value adjusted for False discovery rate (FDR) according to the procedure of Benjamin and Hochberg (57)

**Table S5.**
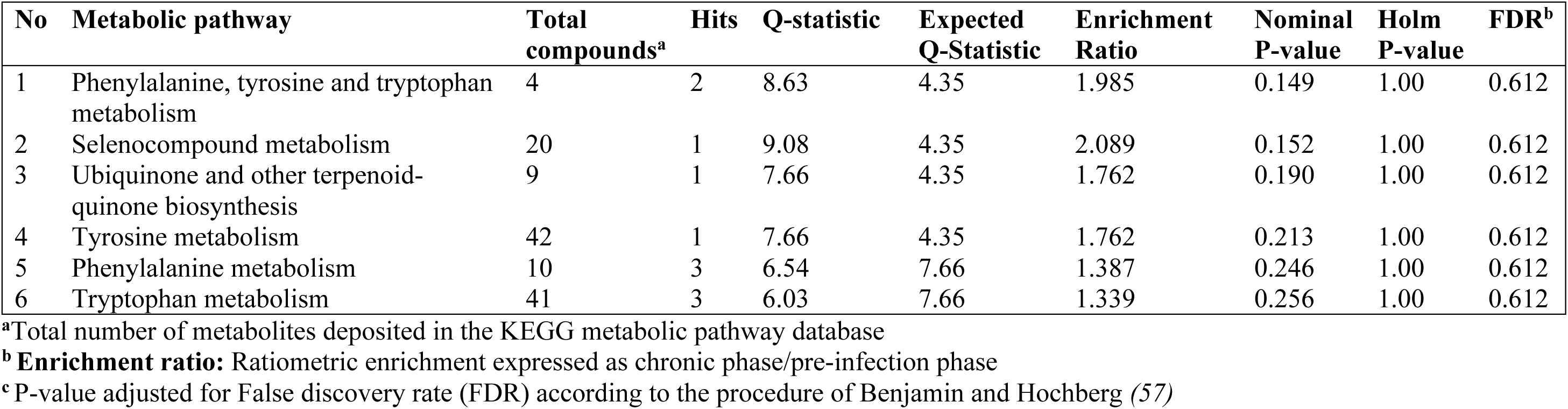
Metabolic Set Enrichment Analysis (MSEA) statistics comparing metabolic pathways enriched during the “chronic phase” of hookworm infection, relative to baseline (i.e. “pre-infection phase”)

**Table S6.** PERMANOVA Analysis identifying “Sex” and “Participant ID” as important covariates to control for in downstream statistical analyses.

| **Factor** | **Df** | **Sum of Squares** | **R<sup>2</sup>** | **F** | **Pr(>F)** |
| --- | --- | --- | --- | --- | --- |
| Sex | 1 | 0.12153 | 0.18997 | 7.9168 | 0.001*** |
| Participant ID | 1 | 0.02956 | 0.04621 | 1.9257 | 0.072 |
| Timepoint | 1 | 0.00865 | 0.01351 | 0.5632 | 0.792 |
| participantID:timepoint | 1 | 0.00412 | 0.00645 | 0.2686 | 0.986 |
| Residual | 31 | 0.47589 | 0.74386 |  |  |
| Total | 35 | 0.363975 | 1 |  |  |

**Significant codes:** $p < 0^{***}$ , $p < 0.001^{**}$ , $p < 0.05^{*}$

**Table S7:** Two-way, repeated-measures Mixed ANOVA analysis, controlling for “sex” and “participant ID” as covariates.

| No | Variables | Effect | DFn | DFd | F | p | p<.05 | ges | p.adj |
| --- | --- | --- | --- | --- | --- | --- | --- | --- | --- |
| 1 | Kynurenine | timepoint | 2 | 33 | 5.108 | 0.012 | * | 0.236 | 0.5076 |
| 2 | Melibiose | timepoint | 2 | 33 | 4.046 | 0.027 | * | 0.197 | 0.5076 |
| 3 | Threonic.acid | timepoint | 2 | 33 | 4.089 | 0.026 | * | 0.199 | 0.5076 |
| 4 | Indoxyl sulfate | timepoint | 2 | 33 | 4.1 | 0.026 | * | 0.199 | 0.5076 |
| 5 | Methoxypsoralen | timepoint | 2 | 33 | 5.503 | 0.009 | * | 0.25 | 0.5076 |

## Data File S1

### Participant Inclusion Criteria

- Participant has provided written informed consent and is willing to comply with all Protocol scheduled visits, laboratory tests, and other trial procedures and in the opinion of the Investigator has a good understanding of the Protocol, the length of the study and the demands of the study.
- Participants will be male and non-pregnant, non-lactating females aged between 18 to 65 years.
- Participants must weigh more than 50kg with a BMI within the 18 – 35kg/m^2^ range
- Participants must understand the procedures involved and agree to participate in the study by giving fully informed, written consent prior to any study assessment.
- Participants must be contactable and available for the duration of the clinical trial.
- If female, has met either of criterion “a or “b” below: **(a) If of non-childbearing potential**, has met 1 of the following – Amenorrhoeic for at least 2 years, or has had a hysterectomy and/or bilateral oophorectomy at least 8 weeks prior to screening, or has had a tubal ligation at least 8 weeks prior to screening. **(b) If of childbearing potential**, must be willing to use the acceptable methods of contraception
- In the opinion of the investigator is in good general health

### Participant Exclusion Criteria

- Current or history of helminth infection (other than *E. vermicularis*).
- Have any finding at screening that in the opinion of the Investigator or medical monitor would compromise the safety of the Participant or affect their ability to adhere to protocol scheduled visits, treatment plan, laboratory tests, and other trial procedures.
- Have participated in any other clinical trial and/or have received an investigational drug or device within 30 days of screening.
- History or current evidence of any of the following: compromised respiratory function (chronic obstructive pulmonary disease, respiratory depression, signs or symptoms of hypoxia at screening); thyroid pathology (unless stabilized and euthyroid for >3 months at the time of screening); hepatitis B, hepatitis C, or human immunodeficiency virus (HIV) infection; evidence of clinically significant chronic cardiac, hepatic or renal disease; psychiatric illness (poorly controlled); seizure disorder or any other chronic health issues that in the opinion of the Investigator would exclude the Participant from the trial.
- Have one of the following laboratory abnormalities: ferritin <20 □g/L, transferrin <2.04 g/L or Hb <120 g/L for females or 130 g/L for males.
- History of severe asthma or other health conditions that may require future steroid use;
- History of substance abuse or current substance abuse that in the opinion of the Investigator would exclude the Participant from the trial.
- History of intolerance, allergy or hypersensitivity to the proposed anthelmintic – mebendazole.
- History of intolerance, allergy or hypersensitivity to the Betadine (iodine) solution used in preparation of *N. americanus* that in the opinion of the Investigator would exclude the participant from the trial.
- History of malignancy of any organ system (other than localized basal cell carcinoma of the skin), treated or untreated, within the past 5 years;
- For female subjects: positive urine pregnancy test at screening.
- Current or past scars, tattoos, or other disruptions of skin integrity at the intended site of larval application.
- Poor venous access making the Participant unable to comply with the safety laboratory testing requirements.
- Probiotic or prebiotic supplementation within 1 month of screening or commencement during the study
- Laxative or gastric motility medication use within 1 month of screening or commencement during the study
- Significant dietary change or weight loss (>5%) within 6 months of screening or during the study
- Smokers or high alcohol consumers

## Data File S2: List of flow cytometry antibodies

Whole Blood Antibodies.

Whole Blood Antibodies
| Antigen | Fluorophore | Clone | #Cat | Supplier |
| --- | --- | --- | --- | --- |
| CD19 | BUV805 | HIB19 | 742007 | BD |
| FceRIa | BV421 | AER-37<br>(CRA-1) | 334624 | Biolegend |
| CD10 | BV480 | HI10a | 746847 | BD |
| CD56 | BV570 | HCD56 | 318330 | Biolegend |
| CD123 | BV605 | 6H6 | 306026 | Biolegend |
| CD11b | BV650 | ICRF44 | 301336 | Biolegend |
| CD15s | BV711 | CSLEX1 | 563910 | BD |
| CD4 | BV750 | SK3 | 344644 | Biolegend |
| Siglec8 | BV786 | 837535 | 747868 | BD |
| HLADR | FITC | G46-6 | 307604 | Biolegend |
| CD3 | AF532 | UCHT1 | 58-0038-42 | Thermofisher |
| CD16 | PerCP | 3G8 | 302030 | Biolegend |
| CXCR4 (CD184) | PE-CF594 | 12G5 | 562389 | BD |
| CD8 | PE-Cy5 | RPA-T8 | 301010 | Biolegend |
| CD203c | APC | NP4D6 | 324610 | Biolegend |
| CD14 | AF700 | HCD14 | 325614 | Biolegend |
| CD15 | APC-Cy7 | W6D3 | 323048 | Biolegend |

PBMC Antibodies.

PBMC Antibodies
| Antigen | Fluorophore | Clone | #Cat | Supplier |
| --- | --- | --- | --- | --- |
| CCR10 | AF488 |  | RDSFAB3478G0100 | R&D Systems |
| CD11c | BV421 | 3.9 | 301628 | Biolegend |
| CD123 | BV605 | 6H6 | 306026 | Biolegend |
| CD127 | BV480 | HIL-7R-M21 | 566101 | BD Biosciences |
| CD14 | PacOrange | TuK4 | MHCD1430 | ThermoFisher |
| CD141 (BDCA3) | PE | AD5-14H12 | 130-113-318 | Miltenyi Biotec |
| CD15 | PE-Cy7 | W6D3 | 323030 | Biolegend |
| CD16 | BUV496 | 3G8 | 612944 | BD Biosciences |
| CD183 (CXCR3) | APC-Cy7 | G025H7 | 353722 | Biolegend |
| CD185 (CXCR5) | BB515 | RF8B2 | 564624 | BD Biosciences |
| CD19 | BV570 | HIB19 | 302236 | Biolegend |
| CD19 | BV711 | Sj25C1 | 563036 | BD Biosciences |
| CD194 (CCR4) | BV605 | L291H4 | 359418 | Biolegend |
| CD196 (CCR6) | BV650 | 11A9 | 563922 | BD Biosciences |
| CD197 (CCR7) | PerCP-Cy5.5 | G043H7 | 353220 | Biolegend |
| CD1c | BV711 | L161 | 331536 | Biolegend |
| CD20 | SB436 | 2H7 | 62-0209-42 | ThermoFisher |
| CD203c | PE-Cy7 | NP4D6 | 324618 | Biolegend |
| CD25 | APC-Fire750 | BC96 | 302642 | Biolegend |
| CD25 | BB515 | 2A3 | 564467 | BD Biosciences |
| CD27 | BUV563 | M-T271 | 741336 | BD Biosciences |
| CD279 (PD-1) | BV480 | EH12.1 | 566112 | BD Biosciences |
| CD28 | BV421 | CD28.2 | 302930 | Biolegend |
| CD294 (CRTH2) | PacBlue | BM16 | 350130 | Biolegend |
| CD3 | BUV496 | UCHT1 | 612940 | BD Biosciences |
| CD3 | PE-Cy7 | UCHT1 | 300420 | Biolegend |
| CD4 | AF532 | RPA-T4 | 58-0049-42 | ThermoFisher |
| CD4 | BV570 | RPA-T4 | 300534 | Biolegend |
| CD45RA | BUV737 | HI100 | 612846 | BD Biosciences |
| CD45RA | BV650 | HI100 | 304136 | Biolegend |
| CD45RO | BV785 | UCHL1 | 304234 | Biolegend |
| CD56 | BUV805 | NCAM16.2 | 749086 | BD Biosciences |
| CD8 | AF532 | RPA-T8 | 58-0088-42 | ThermoFisher |
| CD80 | BV510 | 2D10 | 305234 | Biolegend |
| CD83 | BUV737 | HB15E | 612823 | BD Biosciences |
| CD86 | AF700 | FUN-1 | 561124 | BD Biosciences |
| CTLA-4 | PE-Cy7 | 14D3 | 25-1529-42 | ThermoFisher |
| CTLA-4 | PerCP-eFluor710 | 14D3 | 46-1529-42 | ThermoFisher |
| FceR1a | AF647 | AER-37 | 334614 | Biolegend |
| FoxP3 | AF647 | 259D | 320214 | Biolegend |
| HLA-DR | BUV395 | G46-6 | 564040 | BD Biosciences |
| PLZF | PE-CF594 | R17-809 | 565738 | BD Biosciences |
| TCRgd | BV510 | B1 | 331220 | Biolegend |
| CD1d Tetramer | PE | BS-57 | 53235 | NIH Tetramer Core Facility |
| MRI Tetramer | APC | 5-OP-RU | 53233 | NIH Tetramer Core Facility |
| Zombie NIR Viability Dye |  |  |  | Biolegend |

## Data File S4. Accurate masses and retention times of all annotated metabolites detected in plasma analyzed using negative and positive electrospray ionization modes

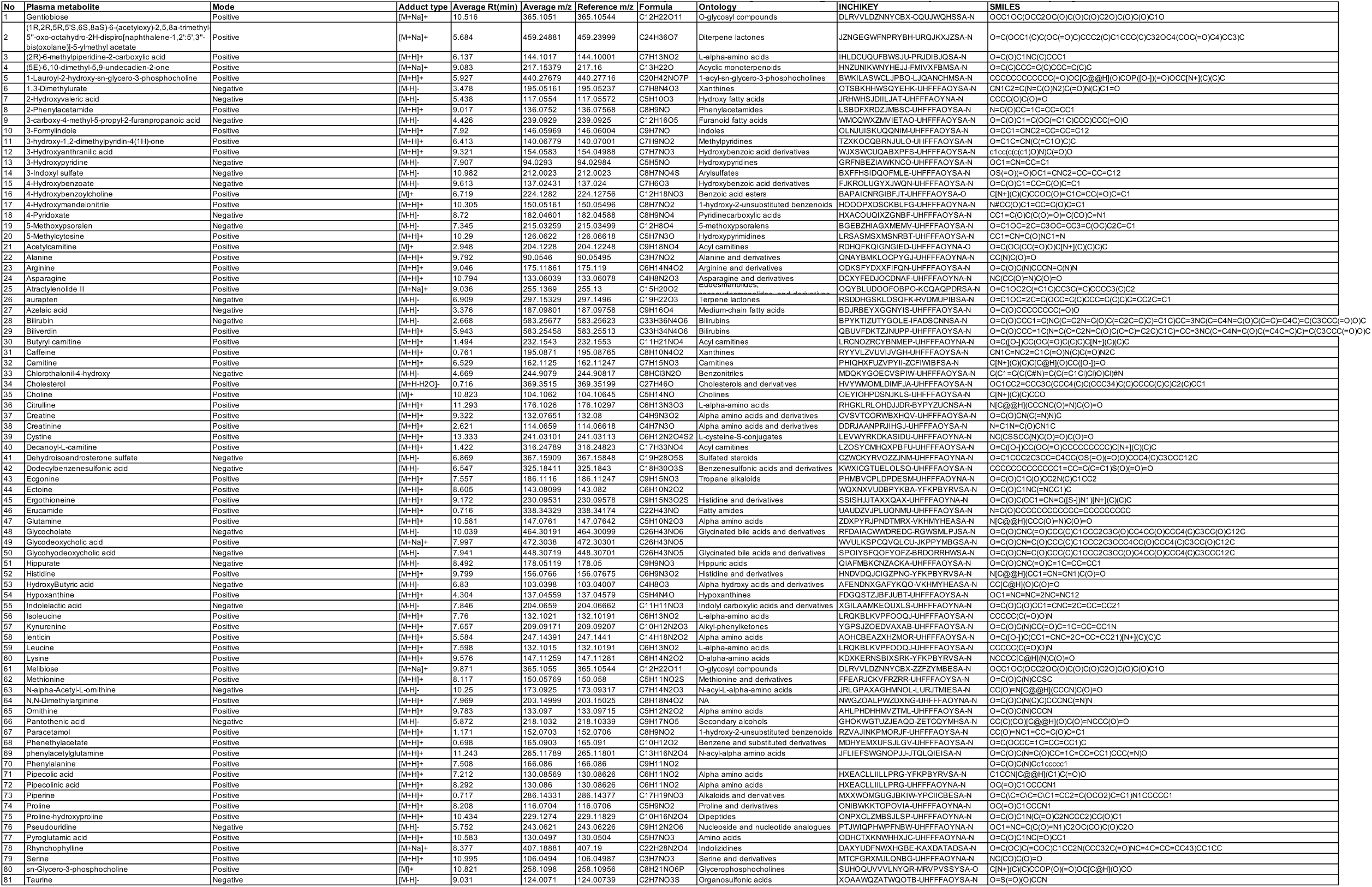

